# Clade I Mpox virus genomic diversity in the Democratic Republic of the Congo, 2018 - 2024: Predominance of Zoonotic Transmission

**DOI:** 10.1101/2024.08.13.24311951

**Authors:** Eddy Kinganda-Lusamaki, Adrienne Amuri-Aziza, Nicolas Fernandez, Jean-Claude Makangara-Cigolo, Catherine Pratt, Emmanuel Hasivirwe Vakaniaki, Nicole A. Hoff, Gradi Luakanda, Prince Akil-Bandali, Sabin Sabiti Nundu, Noella Mulopo-Mukanya, Michel Ngimba, Brigitte Modadra-Madakpa, Ruth Diavita, Princesse Paku, Elisabeth Pukuta-Simbu, Sydney Merritt, Áine O’Toole, Nicola Low, Antoine Nkuba-Ndaye, Hugo Kavunga-Membo, Robert Shongo, Laurens Liesenborghs, Tony Wawina-Bokalanga, Koen Vercauteren, Daniel Mukadi-Bamuleka, Lorenzo Subissi, Jean-Jacques Muyembe, Jason Kindrachuk, Ahidjo Ayouba, Andrew Rambaut, Eric Delaporte, Sofonias Tessema, Eric D’Ortenzio, Anne W. Rimoin, Lisa E. Hensley, Placide Mbala-Kingebeni, Martine Peeters, Steve Ahuka-Mundeke

## Abstract

**Background:** Recent reports raise concerns on the changing epidemiology of mpox in the Democratic Republic of the Congo (DRC), with increasing case counts, sexual contact-mediated clusters, and sustained human-to-human transmission driven by a novel monkeypox virus (MPXV) subclade, clade Ib. However, only a limited number of clade I MPXV genomes have been characterized so far, from a limited number of regions.

**Methods:** We conducted whole genome sequencing of 603 mpox-positive samples that were collected from 581 patients between 2018-2024 in 17 of the 26 provinces of the DRC.

**Results:** Genome coverage was at least 70% for 429/603 (71.1%) samples and near full-length MPXV genomes (>90% coverage) were obtained for 348/603 (57.7%) samples from 337 patients. All newly generated MPXV sequences belonged to clade I, among which 17 were clade Ib strains, all from patients infected in 2024 in the South-Kivu province. The large majority (>95%) of the new strains fall within previously described clade Ia groups and potential new groups have also been observed. The low number of APOBEC3 mutations found among clade Ia suggests that most human mpox cases are probably linked to zoonotic transmissions. Genetically diverse MPXV lineages co-circulate in small geographic areas during the same outbreak suggesting multiple zoonotic introductions over a short period from one or multiple reservoir species. Recent identification of mpox cases in Kinshasa shows that multiple lineages circulate in a large urban center, indicating separate introduction events.

**Conclusion:** The mpox epidemic in the DRC exhibits two distinct patterns. In traditional endemic regions, the epidemic is predominated by zoonotic spill-over events involving clade Ia. Conversely, in the eastern part of the country, the clade Ib outbreak is driven by human-to-human transmission highlighting the need for a coordinated response effort at the national, regional and international levels.

## INTRODUCTION

Mpox, a viral zoonosis caused by Mpox virus (MPXV), is endemic within regions of West and Central Africa. The first case of human mpox was identified in 1970 in the Democratic Republic of the Congo (DRC) (Ladnyj et al, 1972). Since its discovery, cases have been reported in rural and rainforest regions of the Congo Basin in DRC, its neighbouring countries, and in West Africa (Gessain et al 2022). MPXV is divided into two major clades: clade I and II in central and West Africa, respectively (Happi et al, 2022). Cameroon is the only country where clade I and II are known to co-circulate with clades I and II segregated to regions east and west of the country, respectively (Djuici et al ,2024). Clade II is subdivided into clade IIa that contains the viruses from western West Africa, and clade IIb regroups all the sequences from the 2022 global mpox outbreak as well as sequences obtained from Nigeria (Cabanillas et al, 2023; Happi et al, 2022; WHO, 2022a and 2022b). Recent genomic analysis of MPXV strains in eastern DRC demonstrated a distinct subclade within clade I, designated as clade Ib whereas previous phylogenetic studies MPXV from central Africa showed the existence of five main groups (named I to V) in clade I, now referred to as clade Ia (Vakaniaki et al, 2024; Nakazawa et al, 2015; Berthet et al, 2021).

To date, thousands of human cases have been reported in Africa, with the highest burden in DRC (>95% of reported cases), where a national surveillance system has been in place since 2001 (Sklenovská and Van Ranst, 2018; Beer and Rao, 2019; Bunge et al, 2022; Gong et al, 2022; Rimoin et al, 2010). Historically, most mpox outbreaks in Africa have resulted from zoonotic spill-over events followed by limited human-to-human transmission through close contact. Additionally, sporadic travel- or wildlife-associated cases of clade II have been documented in non-endemic regions with an epidemiological link to African foci (Reed et al 2004, Erez et al, 2019; Ng et al, 2019; Vaughan et al, 2018, Mauldin et al, 2022). However, in May 2022, a large mpox outbreak with clade IIb viruses affected 109 non-endemic countries, associated with sexual contact and dense sexual networks among men who have sex with men (Sharif et al, 2023). The outbreak resulted in >90,000 cases worldwide and the declaration of a public health emergency of international concern by the World Health Organization (WHO) (CDC, 2022). Despite a significant decrease in the number of cases worldwide (WHO, 2023), this outbreak is still ongoing (WHO, 2024a; Wertheim et al, 2024).

In DRC, from where most mpox cases have been reported worldwide, the number of suspected mpox cases has doubled in recent years rising from 3,000 cases in 2021 to 5,600 in 2022, and >14,000 in 2023 (WHO 2023c; WHO, 2024b). Since 2023, mpox cases have been reported from an increasing number of provinces, including new foci in large urban areas and transportation hubs, such as Kinshasa (the capital city of the DRC), Bukavu, and Goma in eastern DRC (SGI-Mpox, 2024). Moreover, sexual contact-associated transmission has been reported in multiple regions of the country including Kwango province in the south-west (Kibungu et al., 2024) and a large outbreak in South-Kivu province in the east, a region where only rare cases of mpox were reported before 2023 (McCollum et al, 2015; Vakaniaki et al, 2024). The eastern DRC outbreak was first reported in September 2023 in Kamituga, a mining region, where 297 mpox-confirmed cases and 4 deaths were reported as of the first week of June 2024 (27^th^ Epidemiological week), before spreading to other cities in South-Kivu (Bukavu and Uvira), North-Kivu (Goma) provinces and countries in the eastern region such as Rwanda, Uganda, Kenya and Burundi (INRB, 2024; Oude Munnink et al, 2024; SGI-Mpox, 2024; WHO, 2024b). Signatures of APOBEC3-mediated mutation were observed in these clade Ib strains which suggests human-to-human transmission and clinic-epidemiological data supports that the virus was likely being spread through sexual-contact (Vakaniaki et al, 2024). This observations lead to the declaration of a Public Health Emergency of Continental Security (PHECS) by Africa CDC and a subsequent determination by the WHO Director-General of a Public Health Emergency of International Concern (PHEIC) (WHO 2024d). Genomic sequencing analysis from DRC to date has included a limited number of genomes from only five of 22 affected provinces and the number of MPXV clade I genomes publicly available is insignificant compared to the number of reported cases (McCollum, 2015; Kugelman et al, 2014; Nakazawa et al, 2015, Kibungu et al, 2023; Vakaniaki et al, 2024). In this study, we sequenced and characterized MPXV from samples collected between February 2018 and March 2024 as part of the national surveillance network in order to better document: (i) the genetic diversity of clade I MPXV in DRC and (ii) whether the increase in the number of mpox cases in DRC is due to zoonotic transmission or viral evolution that has been linked to human adaptation and sustained human-to-human transmission.

## MATERIALS AND METHODS

### Ethics statement

Samples were collected as part of national diagnostic and surveillance activities. Authorization for secondary use of the samples for research in this study was obtained from the Ethics Committee of Kinshasa School of Public Health (ESP-UNIKIN, Number ESP/CE/05/2023). The study samples and data were de-identified before genomic and epidemiological analyses.

### Samples

Samples were collected as part of the national Integrated Disease Surveillance and Response (IDSR) program. Cases of mpox were identified by each health zone’s surveillance team according to the ISDR (DRC) and WHO standard definition (WHO, 2022a). Skin lesions (vesicle or scab), blood or oropharyngeal swab, were collected from mpox-suspected individuals. We collected information on age, sex, collection time, time of onset of clinical signs, health zone and province, using the national investigation form. The biological samples were shipped via routine surveillance circuits from health facilities to the provincial level hub and finally from the Provincial Health division to the national reference laboratory (Institut National de Recherche Biomédicale, INRB) in Kinshasa, for PCR confirmation of mpox. Samples from the Eastern part of the country were shipped to the Rodolphe Mérieux-INRB laboratory for PCR confirmation in Goma.

### DNA extraction and molecular testing for mpox diagnosis

Skin lesion swabs were suspended in 1 mL of molecular biology grade PBS 1X prior to inactivation and nucleic acid extraction. DNA was extracted using the QIAamp® DNA Mini kit (Qiagen, Hilden, Germany) or RADI kit (KH Medical, Seoul, South Korea) following the manufacturer’s instructions. Samples were tested for mpox by real-time PCR using either a panel of primers able to detect orthopoxviruses (Kulesh et al, 2004; Li et al, 2010) or with specific MPXV PCR detection assays (RADIPREP kits (http://www.khmedical.co.kr/monkeypox04.php) or Bioperfectus (https://www.bioperfectus.com/ProductDetail/MonkeypoxVirusRealTimePCRKit)) .

### Whole genome sequencing of MPXV

Samples with sufficient volume with positive results for MPXV and by preference with Cycle threshold (Ct) values ≤30 were selected for whole genome sequencing. The Illumina DNA Prep protocol with probe enrichment was used to generate enriched libraries for dual-index paired-end sequencing, using either the comprehensive pan-viral or a custom panel designed for high-consequence viruses, including MPXV (Twist Bioscience, 2021). The enriched libraries were loaded onto an Illumina iSeq100®, Miseq® or Nextseq2000® for 2 x 151 cycles as described previously (Kinganda-Lusamaki et al, 2021). The resulting FASTQ files were analyzed using CZid (https://czid.org) and GeVarLi (https://forge.ird.fr/transvihmi/GeVarLi) for quality control and data cleaning of sequencing reads. Variant calling and genome coverage statistics were then performed, and a consensus genome sequence was generated using the clade I reference genome (Accession number: NC_003310.1). Clade assignment was done using Nextclade–Mpox (all clades, https://clades.nextstrain.org/). All sequences are available in nextstrain.

### Phylogenetic analysis of near-complete MPXV genomes

The new MPXV near-complete genomes from this study were aligned with high-quality and complete MPXV genomes (publicly available and retrieved from NCBI GenBank). These sequences were selected to cover the known clade I diversity (n=98) from DRC and other Central African countries and isolated from humans and animals. Clade II sequences (n=43) were used as outgroup (**Supplementary Table S1)**. Multiple sequence alignment was performed using SQUIRREL (https://github.com/aineniamh/squirrel), which uses Minimap2 to map the genomes against the clade I reference (Accession number NC_003310), aligns them and masks problematic regions (Li H. 2018). The best scalable model, based on aligned data, was obtained using Modelfinder (Kalyaanamoorthy et al., 2017) implemented in IQ-TREE. The phylogeny was inferred by Maximum Likelihood (ML) using IQ-TREE multicore version 2.2.6 (Minh et al., 2020). Branch support was estimated by ultrafast bootstrap with 1000 replicates (Hoang et al., 2018). Trees were formatted and annotated with FigTree, Microreact and iTOL v.5 (Argimón et al, 2016).

### *APOBEC3* analysis of the new MPXV sequences

The new MPXV sequences (with >70% genome coverage) were aligned to the clade I reference genome NC_003310 using Minimap2 v2.17, after exclusion of the 3’ terminal repeat region of the MPXV genome by trimming sequences starting from position 190788 to the end. Additionally, several identified repetitive or low-complexity regions specific to clade I were masked from the alignment. The complete alignment, extraction and masking pipeline can be accessed at http://github.com/aineniamh/squirrel. Subsequently, clade II MPXV genomes dated 1978, 1970 and 1971, were used as outgroups (accession numbers KJ642615, KJ642616 and KJ642617, respectively) in IQ-TREE 2 with the HKY substitution model (Kalyaanamoorthy et al, 2017) to allow estimation of a maximum likelihood tree. Ancestral reconstruction was performed for each internal node of the phylogeny using IQ-TREE 2, allowing mapping of single nucleotide polymorphisms (SNPs) along branches. SNPs were categorized based on their consistency with APOBEC3-editing, assuming that this process induces specific mutations (TC->TT and GA->AA) (O’Toole et al, 2023).

## RESULTS

### Characteristics of patients and samples

A total of 603 samples, collected from 581 patients, were sequenced. The samples were collected between February 2018 and March 2024 as follows; 2018 (n=13), 2020 (n=3), 2021 (n=6), 2022 (n=138; 125 patients), 2023 (n=390; 382 patients) and 2024 (n=53; 52 patients). For a subset of 22 patients, more than one sample type was analysed. Samples originated from 17 of the 26 provinces and 102 of the 519 health zones in DRC (**Figure 1**). The genome coverage was at least 70% for 429 of the 603 (71.1%) samples. High quality genomes (>90% coverage with a minimum depth of 10x) were recovered for 348 (57.7%) samples, representing 337 patients from 14 provinces and 67 health zones. Demographic information including health zone and province locations are provided in **Table 1**. Among the 581 patients, 319 (54.9%) were male, 236 (40.6%) were female and no information was available for 26 (3.7%). The overall median age was 14 years (interquartile reange, IQR: 7-27).

**Figure 1 :**
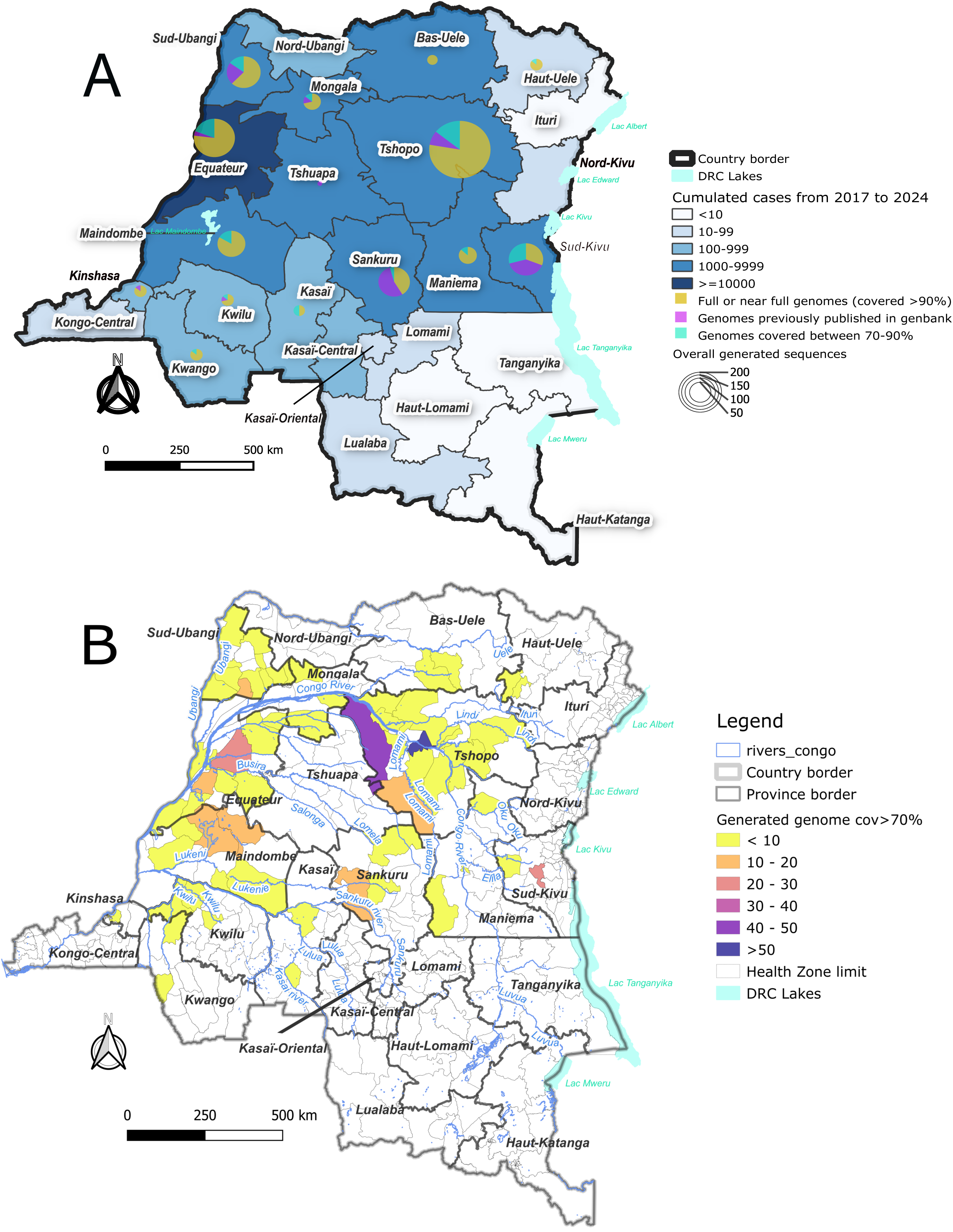
**(1A)** Map with number of MPXV samples sequenced per province between February 2018 and March 2024 represented by size of circles. Different colors illustrate genome coverage from samples in our study (>90%, purple; between 70-90%, brown), and strains available in genbank(pink) from the same provinces. The background map illustrates the number of cumulative suspected mpox cases reported between January 2018 and March 2024 in gradients of blue; **(1B)** Map representing the number of monkeypox virus (MPXV) genomes with more than 70% of genome coverage in the different health zones (HZ) per province in a color gradient of range of samples sequenced.

**Table 1 :** Demographic characteristics of mpox patients with viruses sequenced.

| Province | Number HZ with samples / HZ province | HZ with genome coverage >90%, n (names) | Samples with genome coverage >90%, n | Patients sampled, n | Sex ratio (M/F); missing, n | Age in years, median (IQR) | Collection period, months | genomes, in genbank |
| --- | --- | --- | --- | --- | --- | --- | --- | --- |
| Bas-Uele | 3/11 | 2 (Buta, Titule) | 5 | 8 | 1.7 (5/3) | 29 (12.5-34.5) | 2.5 | - |
| Equateur | 11/18 | 11 (see footnote) | 60 | 91 | 1.1 (43/39); 9 | 11 (5-23) | 24.8 | 3 |
| Haut-Uele | 2/13 | 2 (Isiro, Pawa) | 6 | 10 | 1.5 (6/4) | 25 (10-41) | 21.8 | - |
| Kasaï | 4/18 | 2 (Kamuesha, Ndjoko-punda) | 3 | 7 | 0.8 (3/4) | 18 (9-31) | 0.6 | - |
| Kasaï-Central | 1/26 | - | - | 1 | NA (1/0) | 17 | - | - |
| Kinshasa | 8/34 | 5 (Gombe, Lemba, Limete, Matete, N'sele) | 6 | 12 | 2 (8/4) | 20 (11-23) | 2.3 | 1 |
| Kwango | 4/14 | 2 (Kasongo-Lunda, Kenge) | 6 | 13 | 1.6 (8/5) | 30 (24-40) | 14 | - |
| Kwilu | 7/24 | 4 (Bagata, Kikongo, Masima-nimba, Vanga) | 5 | 8 | 3 (6/2) | 9 (8-20) | 16.5 | 1 |
| Lomami | 1/16 | - | - | 1 | NA (0/1) | 36 | - | - |
| Mai-Ndombe | 7/14 | 5 (Banjow-Moke, Bosobe, Inongo, Kiri, Mushie) | 30 | 58 | 1 (28/29); 1 | 14 (7-30) | 9.2 | - |
| Maniema | 4/18 | 3 (Kindu, Obokote, Tunda) | 13 | 15 | 0.9 (7/8) | 9 (7-21) | 8.3 | - |
| Mongala | 5/12 | 3 (Bongandaganda, Bosomanzi, Bosondjo) | 10 | 16 | 3.7 (11/3); 2 | 10 (4-13) | 11 | 2 |
| North-Ubangi | 2/11 | - | - | 3 | 0.5 (1/2) | 18 (5-35) | - | - |
| Sankuru | 5/16 | 5 (Bena-Dibele, Lodja, Lomela, Ototo, Vanga-kete) | 19 | 26 | 1.4 (15/11) | 15 (4-24) | 67.3 | 25 |
| South-Kivu | 3/34 | 2 (Kadutu, Kamituga) | 17 | 31 | 2 (18/9); 4 | 27 (21-32) | 3 | 22 |
| South-Ubangi | 15/16 | 8 (see footnote) | 33 | 57 | 1.8 (34/19); 4 | 11 (4.5-27) | 14 | 12 |
| Tshopo | 19/23 | 13 (see footnote) | 136 | 224 | 1.3 (125/93); 6 | 13 (7-25) | 73 | 13 |
| Tshuapa | 0/12 | - | - | - | - | - | - | 1 |
| **Total** | **101/330** |  | **348** | **581** | **1.4 (319/236); 26** | **14 (7-27)** | **NA** | **80** |

HZ, health zone; IQR, interquartile range; NA, not applicable;
Equateur health zones: Basankusu, Bikoro, Bolomba, Djombo, Ingende, Lilanga-Bobangi, Lotumbe, Lukolela, Mbandaka, Ntongo and Wangata;
South-Ubangi health zones: Bangabola, Bogose-Nubea, Bokonzi, Boto, Budjala, Bulu, Libenge and Mbaya;
Tshopo health zones: Bafwasende, Basali, Basoko, Bengamisa, Kabondo, Kisangani, Opala, Wanie-Rukula, Yabaondo, Yahuma, Yakusu, Yaleko and Yalimbongo.

Sample types used for sequencing included vesicle (n=267, 44.3%), crust (n=255, 42.3%), blood (n=64, 10.6%) and oropharyngeal swabs (n=17, 2.8%) with the highest genome coverage and sequencing depth obtained from vesicles and crusts. The sequence coverage and quality were highly correlated with viral load with >90% coverage 69% (243/352) of samples with Ct-values <20, 43% (96/221) for Ct-values between 20 and 30, and 11.7% (2/17) with Ct-values >30, independent of the nature of samples (**Supplementary Figure S1**).

### Phylogenetic analysis of MPXV strains: phylogeographic subclusters and spread in urban settings

Analysis of the new MPXV sequences using Nextclade indicated that all sequences belong to clade I. We estimated a maximum likelihood (ML) phylogeny with 348 newly obtained and high quality MPXV genomes (genome coverage >90%) from this study and previously published reference sequences of the different clades and subclades. The 17 new sequences obtained from MPXV-infected individuals in the South-Kivu province in 2024 corresponded to the newly described clade Ib. In contrast, all genomes analysed from elsewhere in the country corresponded to clade Ia (**Figure 2**). According to the previously described clade Ia groups (Berthet et al, 2021; Nakazawa et al, 2015), none of the new sequences belonged to group I which includes only previously reported strains originating from Cameroon and Gabon (**Figure 2a and 2b)**. Group II is the most diverse and widespread across DRC and contains 328 genomes (including 278 (79,9%) of the new genomes and 50 previously published). Group II also includes previously reported sequences from neighbouring countries; i.e. Central African Republic (CAR) (n=10), Sudan (n=2) and Republic of Congo (n=1). Most of the group II sequences in this study came from health zones located around the Congo River or its tributaries such as Ubangi river (North East); Itimbiri, Aruwimi or Uele rivers (North Central); and Kasaï rivers network (South West) (**Figure 2b**). As shown in **Figure 2a**, three main clusters were observed within group II, suggestive of different subgroups. Group III and IV only had MPXV genomes from the DRC (**Figure 2a**). Group III strains circulate in central DRC, mainly Sankuru and Tshopo provinces; group IV mainly circulates across Kwilu, Sankuru, and Maniema provinces, corresponding mainly to the southern savannah area of the country (**Figure 2b**). Group V includes only one previously described isolate from Sankuru District, DRC in 2006 (JX878417). We also identified a new group of 32 sequences that circulates in Tshopo province (**Figure 2a and 2b**). The few previously reported clade Ia genomes from wildlife fall in group I, for the strain obtained from a chimpanzee in Cameroon, and in groups II and III for squirrels (*Funisciurus sp*) and shrews (*Crocidura sp*) from DRC (**Figure 2a**). Interestingly, the six strains from Kinshasa fall into three different lineages from group II and cluster with strains from endemic provinces like Mai-Ndombe and Equateur (**Supplementary Figure S2.**).

**Figure 2 :**
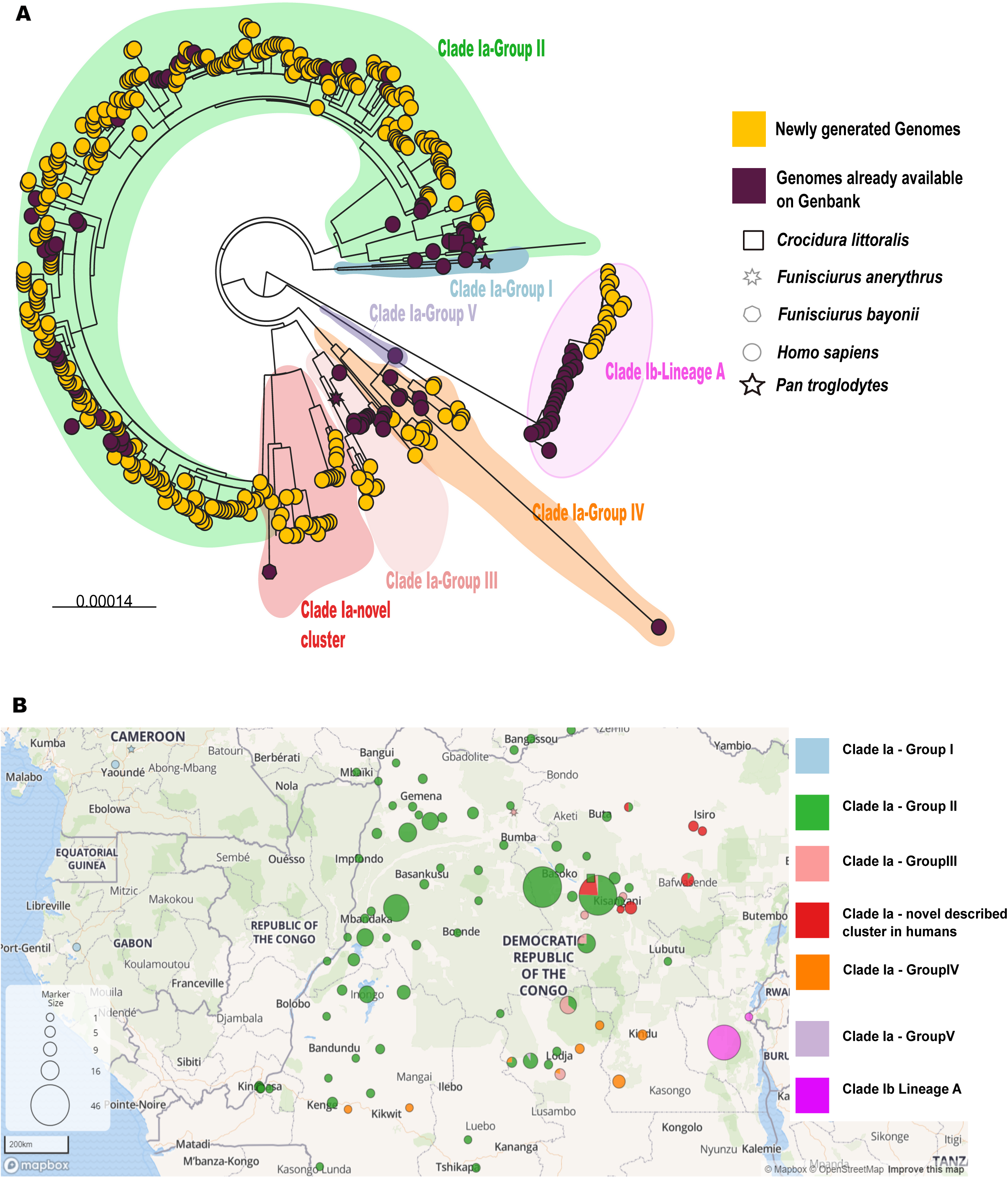
(2A) Phylogenetic tree analysis of the 348 newly obtained MPXV strains from human (yellow dots) and reference strains available in genbank (very dark pink dots for humans and other symbols for animals), as described in Methods. The different clades and groups within clade Ia are highlighted in different colors; clade Ib, pink; clade Ia group I, blue; clade Ia group II, green; clade Ia group III, light orange; clade Ia group IV, dark orange; clade Ia group V; light purple and a potential new group in red; (2B) circles on the map illustrate the geographical location of the different subclades and clade Ia groups in the same colors as in the phylogenetic tree and the numbers are reflected by the size of the circles.

### Co-circulation of different MPXV sub-lineages

In several areas there was co-circulation of different clade Ia groups; in Sankuru province, groups II, IV, and V circulated and given the large geographic spread of group II, co-circulation of group II alongside groups III or IV is seen in other provinces. Importantly, co-circulation of diverse MPXV variants from the same and different groups has also been observed at a smaller scale as shown on the examples in **Figure 3**. In Bikoro health zone (Equateur province), three different viral variants circulated between 22 April and 3 May 2023 (**Figure 3a**). Similar patterns were observed in Yakusu health zone (Tshopo province) where two viral strains circulated between 10 December 2022 and 2 January 2023 (**Figure 3b**) and Bolomba health zone (Equateur province) where two different viral strains circulated between 9 September and 1 October 2022 (**Figure 3c**). In the Inongo health zone (Mai-Ndombe province), three different outbreaks occurred in 2023, each with multiple co-circulating lineages (**Figure 3d**). In contrast, in South Kivu Province, no such diversity was observed and all genomes analysed in this and other studies belonged to a single lineage of clade Ib.

**Figure 3 :**
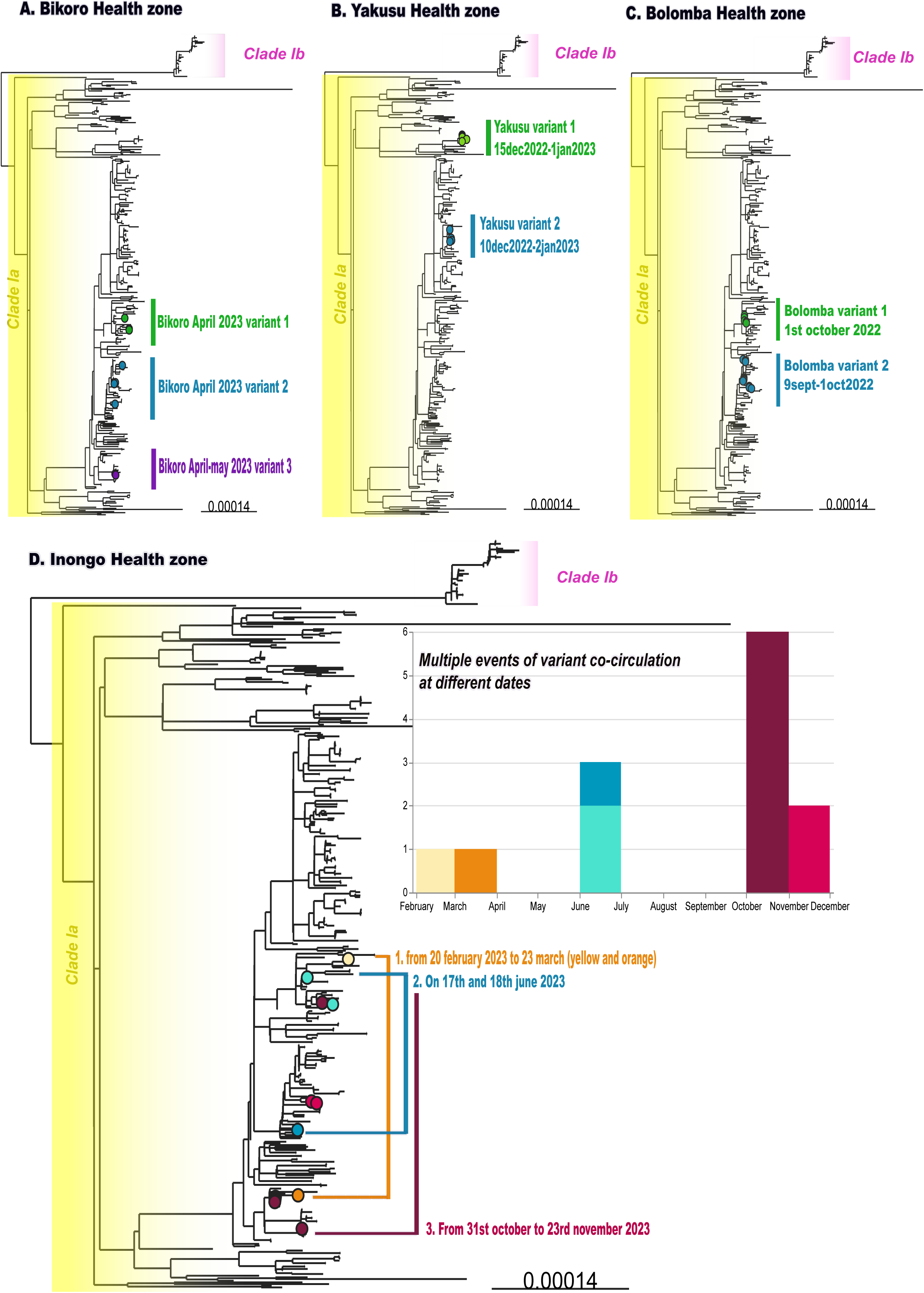
Phylogenetic trees showing examples of health zones where multiple MPXV cocirculate in a same time period, suggesting multiple introductions of viral strains ; (3A) Bikoro health zone in the Equateur Province, three variants cocirculate between April and May 2023 ; (3B) Yakusu health zone in Tshopo province, two variants cocirculate in December 2022 ; (3C) Bolomba health zone in the Equateur province, two variants cocirculate between September and October 2022 in ; (3D) Inongo health zone in the Maindombe Province, three events with cocirculation of multiple variants between February

### Limited APOBEC3 mutations observed among circulating strains

To study the extent of human-to-human transmission, we analysed the different genomes from this study for APOBEC3-related mutations. Here, we included all 429 MPXV sequences with >70% genome coverage and the previously published clade I sequences (**Supplementary Table S1**). Overall, we found very little enrichment of APOBEC3-type mutations in clade Ia sequences. The overall frequency was 10.7% (195/1827) with a ratio (APOBEC3/non-APOBEC3 mutations) of 11.9% (195/1632), ranging from 3.0% to 13.2% (**Table 2**). In-depth analysis of the sequences showed slightly higher rates in some clade Ia sequences; for example, in urban health zones of Kisangani (Tshopo province) and Kindu (Maniema province) and rural areas in Tshopo (Yakusu) and South-Ubangi (Bangabola) provinces (**Supplementary Figure S3**). For clade Ib, 35.9% (23/64) of sequences contained APOBEC3 mutations, with a ratio of 56.1% (23/41).

**Table 2 :** APOBEC3 and other mutations in monkeypox virus (MPXV) subclades Ia and Ib and in the different groups of subclade Ia.

| Clade - group | APOBEC3 | Other | Total | Ratio<br>APOBEC3/other | APOBEC3/Total,<br>% |
| --- | --- | --- | --- | --- | --- |
| **Clade Ia** | **195** | **1632** | **1827** | **0.119** | **10.7%** |
| Clade Ia- Group 1 | 5 | 95 | 100 | 0.053 | 5.0% |
| Clade Ia- Group 2 | 147 | 1112 | 1259 | 0.132 | 11.7% |
| Clade Ia- Group 3 | 11 | 145 | 156 | 0.076 | 7.1% |
| Clade Ia- novel | 28 | 221 | 249 | 0.127 | 11.2% |
| Clade Ia- Group 4 | 1 | 12 | 13 | 0.083 | 7.7% |
| Clade Ia- Group 5 | 1 | 33 | 34 | 0.030 | 2.9% |
| **Overall Clade Ib** | **29** | **111** | **140** | **0.261** | **20.7%** |
| Clade Ib internal<br>branches | 23 | 41 | 64 | 0.561 | 35.9% |

## DISCUSSION

In this report we have characterized the diversity of MPXV in DRC from 2018 to early 2024 and provide insight into the potential drivers underlying the increasing mpox cases. This study generated a total of 348 near-complete (> 90% genome coverage) MPXV genomes from 337 individuals and 14 out of 26 provinces. Almost all (95%) of the new strains belong to clade Ia, in which high genetic diversity is observed. Most new clade Ia strains were in the previously described group II, followed by group III and IV. We also identified new clusters representing potential new groups and sub-groups (Berthet et al, 2021; Nakazawa et al, 2015). In contrast, the new clade Ib strains, all from South Kivu province in 2024, cluster in the single lineage recently described in the South Kivu and North Kivu provinces, with little diversity (Masirika et al, 2024; Vakaniaki et al, 2024, INRB, 2024). Our study highlights two patterns of transmission contributing to the source of human cases: the traditional paradigm of zoonotic spillover with interspecies transmission events and the more recently observed sustained human-to-human transmission driven by sexual contact with an enrichment of APOBEC3-type mutations. The low number of APOBEC3 mutations observed in Clade Ia suggests that most mpox cases have resulted from independent zoonotic introductions into the human population. We also showed that genetically diverse MPXV lineages can co-circulate in small geographic areas during the same outbreak, suggesting multiple zoonotic introductions over a short time span from one or multiple reservoir species. The observation of some clusters from endemic areas with slightly increased numbers of APOBEC3 mutations in MPXV, suggest that there might also be some sustained human transmission in these areas from potential transmission chains that were missed.

The increasing numbers of mpox cases reported from provinces in DRC around the Congo river and its tributaries highlight the potential links between viral spread and human activities around these rivers and reflects possible widespread presence of the animal reservoir for these variants. Human activities could have potentiated MPXV introductions to new health zones and urban areas as illustrated by the introduction of multiple lineages in Kinshasa linked to MPXV strains originating from endemic areas in 2023. These events should be closely monitored as they represent a major threat for additional regional and international dissemination in densely populated areas. This is of particular concern given the increased disease severity associated with clade I MPXV as compared to clade II (Americo et al, 2010; Hutson et al, 2010).

A strength of this study is that we obtained sequences from most of the provinces within DRC that are endemic for mpox with the highest number of mpox cases and provinces, such as South-Kivu with more recent reports of mpox cases. There are also limitations. First, the global picture of MPXV diversity remains incomplete as the 348 MPXV genomes with >90% coverage represent a minority of the suspected mpox cases and only from symptomatic patients seeking health care. More large-scale studies, including genomic surveillance and serosurveys with accompanying epidemiological and clinical data are needed to characterize the extent of the mpox epidemic in humans and the genetic diversity of MPXV strains involved. Secondly, the samples collected and shipped for confirmatory tests were not always from the primary cases (Kinganda-Lusamaki et al, 2022). Thus, more studies are needed for investigating primary cases and a better understanding of viral introduction into the human population.

Our study provides important new information about the genetic diversity of MPXV circulating in DRC. Mpox in DRC is caused predominantly by clade Ia, traditionally associated with zoonotic transmission events. It is thus urgent to better document the nature and extent of animal reservoirs to understand whether the increasing number of cases results from increased direct contacts with putative natural reservoirs, an increase in prevalence of the infection in the animal reservoir, an increase in animal reservoir populations, or a combination of those. The newly described clade Ib was found only in samples from South-Kivu and the limited genetic diversity is compatible with its emergence in 2023 (Vakaniaki et al, 2024). More information is needed about the clinical presentations of mpox caused by different clades and subclades. In the multi-country outbreak of clade IIb, pauci- or asymptomatic cases were documented (Rosenthal, 2024; Debaetselier et al 2022). The clinical presentation of mpox can also differ according to transmission modes, as shown in South-Kivu and North Kivu versus endemic regions in DRC (Vakaniaki et al,2024). The long-term impact of mpox on clinical sequelae, viral dynamics in animal reservoirs and human-to-human transmission, host-immune responses, and persistence of humoral and/or cellular immunity over time need also to be documented.

The recent declaration of a Public Health Emergency of International Concern (PHEIC) (WHO 2024d) has substantial implications for mpox surveillance and for studies of the genomic diversity of MPXV in DRC, regionally and internationally. The global 2022 mpox outbreak highlighted that MPXV is a pathogen of public health concern. The increasing numbers of mpox cases in endemic provinces in DRC and the ongoing outbreak in South-Kivu, which started in a mining city and further disseminated to larger urban areas in DRC such as Bukavu and Goma as well as to four countries in east Africa (Burundi, Kenya, Rwanda and Uganda) previously unaffected by mpox (WHO, 2024c) underscores on the critical need to improve preparedness and coordinate response efforts at the regional and international level. Recent identification of mpox cases in Kinshasa and the presence of multiple lineages in a large urban center also suggest potential regional and international spread with multiple clade I lineages. At the national level, it is urgent to improve access to mpox diagnostic tools, decentralize testing with appropriate materials in health facilities to allow safe and adequate sampling of skin lesions for molecular diagnosis. MPXV genomic surveillance and characterization of MPXV clades and subclades needs to be scaled up, with timely sharing of genome sequences. Nevertheless, the logistical, economic and political challenges for genomic surveillance are as big as the DRC itself, with requirements to access remote areas and deal with security issues in conflict zones. It is time to join efforts to strengthen genomic surveillance in the fight against recurrent mpox epidemics in DRC and worldwide.

## Data Availability

All data produced in the present study are available upon reasonable request to the authors

## ACKNOWLEDGEMENTS

All Provincial Health Division and Health zones, Francois Kasongo, Junior Bulabula, Paul Tshiminyi, Elisabeth Muyamuna, Frida Nkawa, Emmanuel Lokilo, Sifa Kavira, Ola Rilia, Chloé Muswamba, Francisca Muyembe, Emile Muhindo-Milonde, Adèle Kavira-Kamaliro, Zéphanie Paluku-Kalimuli, Jeriel Mubalama-Mufungizi, Tavia Bodisa-Matamu, Raphael Lumembe, Gabriel Kabamba, Michel Kenye, André Citenga, Fiston Cikaya, Meris Matondo, Emile Malembi, Cris Kacita, Thierry Kalonji, Mike R. Wiley, Elise De Vos, Eugene Bangwen, Isabel Brosius, Edyth Parker, CZID Biohub

## FUNDING

This study was supported by Africa CDC Pathogen Genomics Initiative (Africa PGI) (Grants: INV-018278; INV-033857; Saving Lives and Livelihoods program; NU2HGH000077); Agence Française de Développement through the AFROSCREEN project (grant agreement CZZ3209, coordinated by ANRS-MIE Maladies infectieuses émergentes in partnership with Institut de Recherche pour le Développement (IRD) and Pasteur Institute); PANAFPOX project funded by ANRS-MIE ; Belgian Directorate-general Development Cooperation and Humanitarian Aid and the Research Foundation – Flanders (FWO, grant number G096222 N to L.L.); Department of Defense, Defense Threat Reduction Agency, Monkeypox Threat Reduction Network; USDA Non-Assistance Cooperative Agreement #20230048; International mpox Research Consortium (IMReC) through funding from the Canadian Institutes of Health Research and International Development Research Centre (grant no. MRR-184813); Wellcome Trust (Collaborators Award 206298/Z/17/Z, ARTIC network); E.L. received a PhD grant from the French Foreign Office.

## Notes

Conflict of Interest: The authors declare to have no conflict of interest.

### Competing Interest Statement

The authors have declared no competing interest.

### Author Declarations

Ethics Committee of Kinshasa School of Public Health, University of Kinshasa (ESP-UNIKIN, Number ESP/CE/05/2023) gave ethical approval for this work

### Summary of Updates

authors and their affiliation updated, Introduction, figure 2 revised, figures legend updated.

