## Supplementary material for "Clade I Mpox virus genomic diversity in the Democratic Republic of the Congo, 2018 - 2024: Predominance of Zoonotic Transmission": tables

### Supplementary Tables

Supplementary table 1 : Previously published MPXV genomes used in the phylogenetic analysis as contextual genomes with NCBI Genbank accession number (Accession) and related information....... 2
Supplementary table 2 : List of SNP and AA changes within main clades and clusters ......................... 7

### Supplemental tables

*Supplementary table 1 : Previously published MPXV genomes used in the phylogenetic analysis as contextual genomes with NCBI Genbank accession number (Accession) and related information*

| accession | isolate | Host | Country | year | Clade | Authors |
|---|---|---|---|---|---|---|
| DQ011154 | Congo_2003_358 | Homo sapiens | Republic of Congo | 2003 | Ia | Likos et al |
| DQ011155 | Zaire_1979-005 | Homo sapiens | Democratic Republic of the Congo | 1978 | Ia | Likos et al |
| JX878407 | DRC 06-0950 | Homo sapiens | Democratic Republic of the Congo | 2006 | Ia | Kugelman et al |
| JX878408 | DRC 06-0970 | Homo sapiens | Democratic Republic of the Congo | 2006 | Ia | Kugelman et al |
| JX878409 | DRC 06-0999 | Homo sapiens | Democratic Republic of the Congo | 2006 | Ia | Kugelman et al |
| JX878410 | DRC 06-1070 | Homo sapiens | Democratic Republic of the Congo | 2006 | Ia | Kugelman et al |
| JX878411 | DRC 06-1075 | Homo sapiens | Democratic Republic of the Congo | 2006 | Ia | Kugelman et al |
| JX878412 | DRC 06-1076 | Homo sapiens | Democratic Republic of the Congo | 2006 | Ia | Kugelman et al |
| JX878413 | DRC 07-0045 | Homo sapiens | Democratic Republic of the Congo | 2006 | Ia | Kugelman et al |
| JX878414 | DRC 07-0046 | Homo sapiens | Democratic Republic of the Congo | 2006 | Ia | Kugelman et al |
| JX878415 | DRC 07-0092 | Homo sapiens | Democratic Republic of the Congo | 2006 | Ia | Kugelman et al |
| JX878416 | DRC 07-0093 | Homo sapiens | Democratic Republic of the Congo | 2006 | Ia | Kugelman et al |
| JX878417 | DRC 07-0104 | Homo sapiens | Democratic Republic of the Congo | 2006 | Ia | Kugelman et al |
| JX878418 | DRC 07-0120 | Homo sapiens | Democratic Republic of the Congo | 2007 | Ia | Kugelman et al |
| JX878419 | DRC 07-0275 | Homo sapiens | Democratic Republic of the Congo | 2007 | Ia | Kugelman et al |
| JX878420 | DRC 07-0283 | Homo sapiens | Democratic Republic of the Congo | 2007 | Ia | Kugelman et al |
| JX878421 | DRC 07-0286 | Homo sapiens | Democratic Republic of the Congo | 2007 | Ia | Kugelman et al |
| JX878422 | DRC 07-0287 | Homo sapiens | Democratic Republic of the Congo | 2007 | Ia | Kugelman et al |
| JX878423 | DRC 07-0337 | Homo sapiens | Democratic Republic of the Congo | 2007 | Ia | Kugelman et al |
| JX878424 | DRC 07-0338 | Homo sapiens | Democratic Republic of the Congo | 2007 | Ia | Kugelman et al |
| JX878425 | DRC 07-0354 | Homo sapiens | Democratic Republic of the Congo | 2007 | Ia | Kugelman et al |
| JX878426 | DRC 07-0450 | Homo sapiens | Democratic Republic of the Congo | 2007 | Ia | Kugelman et al |
| JX878427 | DRC 07-0480 | Homo sapiens | Democratic Republic of the Congo | 2007 | Ia | Kugelman et al |
| JX878428 | DRC 07-0514 | Homo sapiens | Democratic Republic of the Congo | 2007 | Ia | Kugelman et al |

| accession | isolate | Host | Country | year | Clade | Authors |
|---|---|---|---|---|---|---|
| JX878429 | DRC 07-0662 | Homo sapiens | Democratic Republic of the Congo | 2007 | Ia | Kugelman et al |
| KC257459 | | Homo sapiens | Sudan | 2005 | Ia | Nakazawa,Y., et al. |
| KC257460 | KC257460 | Homo sapiens | Democratic Republic of the Congo | 1985 | Ia | Nakazawa et al |
| KJ642612 | KJ642612 | Homo sapiens | Democratic Republic of the Congo | 1986 | Ia | Nakazawa et al |
| KJ642613 | KJ642613 | Homo sapiens | Democratic Republic of the Congo | 1970 | Ia | Nakazawa et al |
| KJ642618 | KJ642618 | Homo sapiens | Cameroon | 1989 | Ia | Nakazawa et al |
| KJ642619 | KJ642619 | Homo sapiens | Gabon | 1987 | Ia | Nakazawa et al |
| KP849469 | Boende_DRC_2008 | Homo sapiens | Democratic Republic of the Congo | 2008 | Ia | Nakazawa et al |
| KP849471 | Yambuku_DRC_1985 | Funisciurus anerythrus | Democratic Republic of the Congo | 1985 | Ia | Nakazawa et al |
| MN702444 | MN702444 | Homo sapiens | Central African Republic | 2017 | Ia | Selekon et al |
| MN702445 | MN702445 | Homo sapiens | Central African Republic | 2017 | Ia | Selekon et al |
| MN702446 | MN702446 | Homo sapiens | Central African Republic | 2018 | Ia | Selekon et al |
| MN702447 | MN702447 | Homo sapiens | Central African Republic | 2018 | Ia | Selekon et al |
| MN702448 | MN702448 | Homo sapiens | Central African Republic | 2018 | Ia | Selekon et al |
| MN702449 | MN702449 | Homo sapiens | Central African Republic | 2016 | Ia | Selekon et al |
| MN702450 | MN702450 | Homo sapiens | Central African Republic | 2016 | Ia | Selekon et al |
| MN702451 | MN702451 | Homo sapiens | Central African Republic | 2017 | Ia | Selekon et al |
| MN702452 | MN702452 | Homo sapiens | Central African Republic | 2010 | Ia | Selekon et al |
| MN702453 | MN702453 | Homo sapiens | Central African Republic | 2001 | Ia | Selekon et al |
| MT724770 | MT724770 | Crocidura littoralis | Democratic Republic of the Congo | 2014 | Ia | Marien et al |
| MT724771 | MT724771 | Funisciurus bayonii | Democratic Republic of the Congo | 2014 | Ia | Marien et al |
| MT724772 | MT724772 | Funisciurus anerythrus | Democratic Republic of the Congo | 2014 | Ia | Marien et al |
| NC_003310 | | Homo sapiens | Democratic Republic of the Congo | 1996 | Ia | Senkevich,T.G., et al. |
| OP498046 | BNITM-Gabon1988 | Homo sapiens | Gabon | 1988 | Ia | Emmerich et al |
| OQ621553 | MPXV/Sudan/2022/RKI630 | Homo sapiens | Sudan | 2022 | Ia | Brinkmann,A., et al. |
| OQ729808 | MPXV/COD/2023/172V | Homo sapiens | Democratic Republic of the Congo | 2022 | Ia | Lee et al |
| OR943698 | MPXV_CMR_2016_Chimp | Pan troglodytes | Cameroon | 2016 | Ia | Brien et al |
| PP601182 | 23MPX1740C | Homo sapiens | Democratic Republic of the Congo | 2023 | Ia | Vakaniaki et al |
| PP601183 | 23MPX1766C | Homo sapiens | Democratic Republic of the Congo | 2023 | Ia | Vakaniaki et al |

| accession | isolate | Host | Country | year | Clade | Authors |
|---|---|---|---|---|---|---|
| PP601184 | 23MPX1769C | Homo sapiens | Democratic Republic of the Congo | 2023 | Ia | Vakaniaki et al |
| PP601185 | 23MPX1786C | Homo sapiens | Democratic Republic of the Congo | 2023 | Ia | Vakaniaki et al |
| PP601186 | 23MPX1793V | Homo sapiens | Democratic Republic of the Congo | 2023 | Ia | Vakaniaki et al |
| PP601187 | 23MPX1806V | Homo sapiens | Democratic Republic of the Congo | 2023 | Ia | Vakaniaki et al |
| PP601188 | 24MPX0008V | Homo sapiens | Democratic Republic of the Congo | 2023 | Ia | Vakaniaki et al |
| PP601189 | 24MPX0009C | Homo sapiens | Democratic Republic of the Congo | 2023 | Ia | Vakaniaki et al |
| PP601190 | 24MPX0012C | Homo sapiens | Democratic Republic of the Congo | 2023 | Ia | Vakaniaki et al |
| PP601191 | 24MPX0014C | Homo sapiens | Democratic Republic of the Congo | 2023 | Ia | Vakaniaki et al |
| PP601192 | 24MPX0018V | Homo sapiens | Democratic Republic of the Congo | 2024 | Ia | Vakaniaki et al |
| PP601193 | 24MPX0024C | Homo sapiens | Democratic Republic of the Congo | 2023 | Ia | Vakaniaki et al |
| PP601194 | 24MPX0025V | Homo sapiens | Democratic Republic of the Congo | 2023 | Ia | Vakaniaki et al |
| PP601195 | 24MPX0026V | Homo sapiens | Democratic Republic of the Congo | 2023 | Ia | Vakaniaki et al |
| PP601196 | 24MPX0037V | Homo sapiens | Democratic Republic of the Congo | 2024 | Ia | Vakaniaki et al |
| PP601197 | 24MPX0038V | Homo sapiens | Democratic Republic of the Congo | 2024 | Ia | Vakaniaki et al |
| PP601198 | 24MPX0041V | Homo sapiens | Democratic Republic of the Congo | 2024 | Ia | Vakaniaki et al |
| PP601199 | 24MPX0164V | Homo sapiens | Democratic Republic of the Congo | 2024 | Ia | Vakaniaki et al |
| PP601200 | 24MPX0166C | Homo sapiens | Democratic Republic of the Congo | 2024 | Ia | Vakaniaki et al |
| PP601201 | 24MPX0168V | Homo sapiens | Democratic Republic of the Congo | 2024 | Ia | Vakaniaki et al |
| PP601202 | 24MPX0169C | Homo sapiens | Democratic Republic of the Congo | 2024 | Ia | Vakaniaki et al |
| PP601203 | 24MPX0174C | Homo sapiens | Democratic Republic of the Congo | 2024 | Ia | Vakaniaki et al |
| PP601204 | 24MPX0175V | Homo sapiens | Democratic Republic of the Congo | 2024 | Ia | Vakaniaki et al |
| PP601205 | 24MPX0188V | Homo sapiens | Democratic Republic of the Congo | 2024 | Ia | Vakaniaki et al |
| PP601206 | 24MPX0194V | Homo sapiens | Democratic Republic of the Congo | 2024 | Ia | Vakaniaki et al |
| PP601207 | 24MPX0198V | Homo sapiens | Democratic Republic of the Congo | 2024 | Ib | Vakaniaki et al |
| PP601208 | 24MPX0201V | Homo sapiens | Democratic Republic of the Congo | 2024 | Ib | Vakaniaki et al |
| PP601209 | 24MPX0203V | Homo sapiens | Democratic Republic of the Congo | 2024 | Ib | Vakaniaki et al |
| PP601210 | 24MPX0205V | Homo sapiens | Democratic Republic of the Congo | 2024 | Ib | Vakaniaki et al |
| PP601211 | 24MPX0206V | Homo sapiens | Democratic Republic of the Congo | 2024 | Ib | Vakaniaki et al |
| PP601212 | 24MPX0207Or | Homo sapiens | Democratic Republic of the Congo | 2024 | Ib | Vakaniaki et al |
| PP601213 | 24MPX0209V | Homo sapiens | Democratic Republic of the Congo | 2024 | Ib | Vakaniaki et al |

| accession | isolate | Host | Country | year | Clade | Authors |
|---|---|---|---|---|---|---|
| PP601214 | 24MPX0214V | Homo sapiens | Democratic Republic of the Congo | 2024 | Ib | Vakaniaki et al |
| PP601215 | 24MPX0217V | Homo sapiens | Democratic Republic of the Congo | 2024 | Ib | Vakaniaki et al |
| PP601216 | 24MPX0220V | Homo sapiens | Democratic Republic of the Congo | 2024 | Ib | Vakaniaki et al |
| PP601217 | 24MPX0221V | Homo sapiens | Democratic Republic of the Congo | 2024 | Ib | Vakaniaki et al |
| PP601218 | 24MPX0223C | Homo sapiens | Democratic Republic of the Congo | 2023 | Ib | Vakaniaki et al |
| PP601219 | 24MPX0224C | Homo sapiens | Democratic Republic of the Congo | 2023 | Ib | Vakaniaki et al |
| PP601220 | 24MPX0226V | Homo sapiens | Democratic Republic of the Congo | 2023 | Ib | Vakaniaki et al |
| PP601221 | 24MPX0228V | Homo sapiens | Democratic Republic of the Congo | 2023 | Ib | Vakaniaki et al |
| PP601222 | 24MPX0230C | Homo sapiens | Democratic Republic of the Congo | 2023 | Ib | Vakaniaki et al |
| PP601223 | 24MPX0231V | Homo sapiens | Democratic Republic of the Congo | 2023 | Ib | Vakaniaki et al |
| PP601224 | 24MPX0239V | Homo sapiens | Democratic Republic of the Congo | 2024 | Ib | Vakaniaki et al |
| PP601225 | 24MPX0240V | Homo sapiens | Democratic Republic of the Congo | 2024 | Ib | Vakaniaki et al |
| PP601226 | 24MPX0242V | Homo sapiens | Democratic Republic of the Congo | 2024 | Ib | Vakaniaki et al |
| PP601227 | RDC-NKV-GOM-MPOX-004 | Homo sapiens | Democratic Republic of the Congo | 2023 | Ib | Vakaniaki et al |
| PP601228 | RDC-NKV-GOM-MPOX-010 | Homo sapiens | Democratic Republic of the Congo | 2023 | Ib | Vakaniaki et al |
| AY603973 | | Cynomolgus monkey | Danemark | 1961 | IIa | Chen N et al |
| AY741551 | Sierra Leone | Homo sapiens | Sierra Leone | 1970 | IIa | Chen N et al |
| AY753185 | | Cynomolgus monkey | Danemark | 1958 | IIa | Chen N et al |
| KJ642615 | | Homo sapiens | Nigeria | 1978 | IIa | Nakazawa et al |
| KJ642616 | | Pan troglodytes | France | 1968 | IIa | Nakazawa et al |
| KJ642617 | | Homo sapiens | Nigeria | 1971 | IIa | Nakazawa et al |
| KP849470 | Cote d'Ivoire_1971 | Homo sapiens | Côte d'Ivoire | 1971 | IIa | Nakazawa et al |
| MN346690 | MPXV_TNP_2017_North_Bic | Pan troglodytes | Côte d'Ivoire | 2017 | IIa | Patrono L.V et al |
| MN346692 | MPXV_TNP_2017_North_Mama | Pan troglodytes | Côte d'Ivoire | 2017 | IIa | Patrono L.V et al |
| MN346693 | MPXV_TNP_2017_North_Ponan | Pan troglodytes | Côte d'Ivoire | 2017 | IIa | Patrono L.V et al |
| MN346694 | MPXV_TNP_2017_North_Saro | Pan troglodytes | Côte d'Ivoire | 2017 | IIa | Patrono L.V et al |
| MN346695 | MPXV_TNP_2017_North_Sidonie | Pan troglodytes | Côte d'Ivoire | 2017 | IIa | Patrono L.V et al |
| MN346696 | MPXV_TNP_2017_North_Surprise_1 | Pan troglodytes | Côte d'Ivoire | 2017 | IIa | Patrono L.V et al |
| MN346697 | MPXV_TNP_2017_North_Surprise_2 | Pan troglodytes | Côte d'Ivoire | 2017 | IIa | Patrono L.V et al |
| MN346698 | MPXV_TNP_2017_South_Pushkin | Pan troglodytes | Côte d'Ivoire | 2017 | IIa | Patrono L.V et al |

| accession | isolate | Host | Country | year | Clade | Authors |
|---|---|---|---|---|---|---|
| MN346699 | MPXV_TNP_2017_South_Ravel_1 | Pan troglodytes | Côte d'Ivoire | 2017 | IIa | Patrono L.V et al |
| MN346700 | MPXV_TNP_2017_South_Ravel_2 | Pan troglodytes | Côte d'Ivoire | 2017 | IIa | Patrono L.V et al |
| MN346701 | MPXV_TNP_2017_South_Woodstock | Pan troglodytes | Côte d'Ivoire | 2017 | IIa | Patrono L.V et al |
| MN346702 | MPXV_TNP_2018_East_Paddy | Pan troglodytes | Côte d'Ivoire | 2018 | IIa | Patrono L.V et al |
| MN346703 | MPXV_TNP_2018_East_Placali | Pan troglodytes | Côte d'Ivoire | 2018 | IIa | Patrono L.V et al |
| MT903346 | MPXV-USA2003_099_Gambian_Rat | Cricetomys gambianus | USA | 2003 | IIa | Mauldin M et al |
| MT903347 | MPXV-USA2003_099_Dormouse | Gliridae | USA | 2003 | IIa | Mauldin M et al |
| MT903348 | MPXV-USA2003_099_Rope_Squirrel | Funisciurus | USA | 2003 | IIa | Mauldin M et al |
| MT903339 | MPXV-M3021_Delta | Homo sapiens | Nigeria | 2018 | IIb | Mauldin M et al |
| MT903340 | MPXV-M5312_HM12_Rivers | Homo sapiens | Nigeria | 2018 | IIb | Mauldin M et al |
| MT903342 | MPXV-Singapore | Homo sapiens | Singapore | 2018 | IIb | Mauldin M et al |
| MT903343 | MPXV-UK_P1 | Homo sapiens | UK | 2018 | IIb | Mauldin M et al |
| NC_063383 | MPXV-M5312_HM12_Rivers | Homo sapiens | Nigeria | 2018 | IIb | Mauldin M et al |
| ON622712 | MPX/UZ_REGA_1/Belgium/2022 | Homo sapiens | Belgium | 2022 | IIb | Vanmechelen et al |
| ON918611 | NICD-SVPL223 | Homo sapiens | South-Africa | 2022 | IIb | Chan et al |
| ON927248 | NICD-SVPL232 | Homo sapiens | South-Africa | 2022 | IIb | Chan et al |
| OP422631 | MPXV-SEN0021 | Homo sapiens | Benin | 2022 | IIb | Martin et al |
| OP422632 | MPXV-SEN0022 | Homo sapiens | Benin | 2022 | IIb | Martin et al |
| OP535312 | MPXV_Nigeria_2018_5222 | Homo sapiens | Nigeria | 2018 | IIb | Nmaemeka et al |
| OP535317 | MPXV_Nigeria_2018_5194 | Homo sapiens | Nigeria | 2018 | IIb | Nmaemeka et al |
| OP535320 | MPXV_Nigeria_2017_3008 | Homo sapiens | Nigeria | 2017 | IIb | Nmaemeka et al |
| OP535322 | MPXV_Nigeria_2017_2958 | Homo sapiens | Nigeria | 2017 | IIb | Nmaemeka et al |
| OP535324 | MPXV_Nigeria_2018_5302 | Homo sapiens | Nigeria | 2018 | IIb | Nmaemeka et al |
| OP535325 | MPXV_Nigeria_2018_5316 | Homo sapiens | Nigeria | 2018 | IIb | Nmaemeka et al |
| OP535327 | MPXV_Nigeria_2018_5307 | Homo sapiens | Nigeria | 2018 | IIb | Nmaemeka et al |
| OP535337 | MPXV_Nigeria_2017_2945 | Homo sapiens | Nigeria | 2017 | IIb | Nmaemeka et al |
| PP338787 | MPXV/Human/USA/CA-LACPHL-MA00577/2024 | Homo sapiens | USA | 2024 | IIb | McCann et al |
| PP496048 | hMpxV/Philippines/RITM-002/2022 | Homo sapiens | Philippines | 2022 | IIb | Balingit et al |

*Supplementary table 2 : List of SNP and AA changes within main clades and clusters*

| site | direction | snp | dimer | apobec | parent codon | parent aa | child codon | child aa | mutation_category | prediction | homoplasy | Cluster | Remarks | gene-OPG |
|---|---|---|---|---|---|---|---|---|---|---|---|---|---|---|
| 241 | NA | A->C | | No | NA | NA | NA | NA | intergenic | NA | Yes | Clade Ia | | NA |
| 172022 | NA | A->G | | No | NA | NA | NA | NA | intergenic | NA | No | Clade Ia | | NA |
| 178933 | NA | A->G | | No | NA | NA | NA | NA | intergenic | NA | Yes | Clade Ia | | NA |
| 187600 | NA | G->A | GC | No | NA | NA | NA | NA | intergenic | NA | No | Clade Ia | | NA |
| 37379 | reverse | C->A | | No | CGG | R | CTG | L | nonsynonymous | moderately radical | No | Clade Ia | | OPG056 |
| 127648 | forward | A->G | | No | ACC | T | GCC | A | nonsynonymous | moderately conservative | No | Clade Ia | | OPG149 |
| 140473 | reverse | T->C | | No | AGT | S | GGT | G | nonsynonymous | moderately conservative | No | Clade Ia | | OPG160 |
| 152010 | forward | G->A | GC | No | CGC | R | CAC | H | nonsynonymous | conservative | No | Clade Ia | | OPG178 |
| 1899 | reverse | G->A | GA | Yes | GTC | V | GTT | V | synonymous | NA | No | Clade Ia group I | | OPG002 |
| 97172 | forward | G->A | GG | No | GAG | E | GAA | E | synonymous | NA | No | Clade Ia group I | | OPG117 |
| 107603 | reverse | G->T | | No | GTC | V | GTA | V | synonymous | NA | No | Clade Ia group I | | OPG125 |
| 120895 | reverse | C->T | GC | No | ATG | M | ATA | I | nonsynonymous | conservative | No | Clade Ia group I | | OPG138 |
| 123589 | reverse | C->T | GC | No | GCT | A | ACT | T | nonsynonymous | moderately conservative | No | Clade Ia group I | | OPG144 |
| 146465 | reverse | A->G | | No | TTA | L | CTA | L | synonymous | NA | No | Clade Ia group I | | OPG170 |
| 188518 | NA | G->A | GC | No | NA | NA | NA | NA | intergenic | NA | No | Clade Ia excepted JX874717 | | NA |
| 1030 | reverse | G->A | GA | Yes | TCA | S | TTA | L | nonsynonymous | moderately radical | No | Clade Ia excepted JX874717 | | OPG001 |
| 16196 | reverse | G->T | | No | CTA | L | ATA | I | nonsynonymous | conservative | No | Clade Ia excepted JX874717 | | OPG027 |
| 74384 | forward | C->T | TC | Yes | ATC | I | ATT | I | synonymous | NA | No | Clade Ia excepted JX874717 | | OPG094 |
| 186631 | forward | G->T | | No | GAG | E | GAT | D | nonsynonymous | conservative | No | Clade Ia excepted JX874717 | | OPG210 |
| 169500 | NA | G->A | GT | No | NA | NA | NA | NA | intergenic | NA | Yes | Clade Ia, groups II and III | | NA |
| 169501 | NA | T->A | | No | NA | NA | NA | NA | intergenic | NA | Yes | Clade Ia, groups II and III | | NA |

| site | direction | snp | dimer | apobec | parent codon | parent aa | child codon | child aa | mutation_category | prediction | homoplasy | Cluster | Remarks | gene-OPG |
|---|---|---|---|---|---|---|---|---|---|---|---|---|---|---|
| 132943 | NA | C->T | GC | No | NA | NA | NA | NA | intergenic | NA | No | Clade Ia, group II | | NA |
| 1355 | reverse | G->T | | No | CTA | L | ATA | I | nonsynonymous | conservative | No | Clade Ia, group II | | OPG001 |
| 73148 | forward | G->A | GA | Yes | GAG | E | AAG | K | nonsynonymous | moderately conservative | No | Clade Ia, group II | | OPG093 |
| 137411 | reverse | G->A | GC | No | CGT | R | TGT | C | nonsynonymous | radical | No | Clade Ia, group II | | OPG153 |
| 159492 | forward | T->C | | No | TTT | F | CTT | L | nonsynonymous | conservative | No | Clade Ia, group II | | OPG185 |
| 7732 | forward | C->A | | No | CCA | P | ACA | T | nonsynonymous | conservative | No | Clade Ia group II, III and novel | | OPG019 |
| 8973 | NA | C->A | | No | NA | NA | NA | NA | intergenic | NA | No | Clade Ia, group II branch 1 | | NA |
| 20627 | reverse | G->A | GA | Yes | TCC | S | TTC | F | nonsynonymous | radical | No | Clade Ia, group II branch 1 | | OPG036 |
| 58283 | reverse | A->G | | No | GTT | V | GTC | V | synonymous | NA | No | Clade Ia, group II branch 1 | | OPG077 |
| 83797 | forward | C->T | GC | No | AGC | S | AGT | S | synonymous | NA | No | Clade Ia, group II branch 1 | | OPG105 |
| 97065 | forward | C->T | GC | No | CAT | H | TAT | Y | nonsynonymous | moderately conservative | No | Clade Ia, group II branch 1 | | OPG117 |
| 143163 | forward | C->T | CC | No | GCC | A | GCT | A | synonymous | NA | No | Clade Ia, group II branch 1 | | OPG164 |
| 149978 | forward | G->A | GT | No | ATG | M | ATA | I | nonsynonymous | conservative | Yes | Clade Ia, group II branch 1 | | OPG176 |
| 620 | NA | A->G | | No | NA | NA | NA | NA | intergenic | NA | Yes | Clade Ia, group II branch2 to KJ642613 | | NA |
| 7445 | NA | G->A | GT | No | NA | NA | NA | NA | intergenic | NA | No | Clade Ia, group II branch2 to KJ642613 | | NA |
| 8224 | NA | C->T | GC | No | NA | NA | NA | NA | intergenic | NA | Yes | Clade Ia, group II branch2 to KJ642613 | | NA |
| 13359 | NA | G->A | GT | No | NA | NA | NA | NA | intergenic | NA | No | Clade Ia, group II branch2 to KJ642613 | | NA |
| 33659 | NA | C->T | GC | No | NA | NA | NA | NA | intergenic | NA | No | Clade Ia, group II branch2 to KJ642613 | | NA |
| 135781 | NA | G->A | GC | No | NA | NA | NA | NA | intergenic | NA | Yes | Clade Ia, group II branch2 to KJ642613 | | NA |
| 178921 | NA | T->A | | No | NA | NA | NA | NA | intergenic | NA | Yes | Clade Ia, group II branch2 to KJ642613 | | NA |

| site | direction | snp | dimer | apobec | parent codon | parent aa | child codon | child aa | mutation_category | prediction | homoplasy | Cluster Remarks | gene-OPG |
|---|---|---|---|---|---|---|---|---|---|---|---|---|---|
| 60686 | reverse | G->A | GA | Yes | CTC | L | CTT | L | synonymous | NA | No | Clade Ia, group II branch2 to KJ642613 | OPG080 |
| 65424 | forward | G->A | GC | No | CGC | R | CAC | H | nonsynonymous | conservative | No | Clade Ia, group II branch2 to KJ642613 | OPG084 |
| 66885 | reverse | C->T | AC | No | TGT | C | TAT | Y | nonsynonymous | radical | No | Clade Ia, group II branch2 to KJ642613 | OPG085 |
| 68972 | forward | G->T | | No | ACG | T | ACT | T | synonymous | NA | No | Clade Ia, group II branch2 to KJ642613 | OPG087 |
| 81642 | forward | C->T | AC | No | ACG | T | ATG | M | nonsynonymous | moderately conservative | No | Clade Ia, group II branch2 to KJ642613 | OPG105 |
| 113802 | reverse | C->T | TC | Yes | GAG | E | AAG | K | nonsynonymous | moderately conservative | No | Clade Ia, group II branch2 to KJ642613 | OPG133 |
| 146163 | reverse | G->T | | No | TCC | S | TCA | S | synonymous | NA | No | Clade Ia, group II branch2 to KJ642613 | OPG170 |
| 12677 | reverse | G->A | GC | No | CTC | L | TTC | F | nonsynonymous | conservative | No | Clade Ia, group II branch 2 | OPG023 |
| 37094 | NA | G->A | GG | No | NA | NA | NA | NA | intergenic | NA | No | Clade Ia, group II to branch 2 excepted KJ642613 | NA |
| 156635 | NA | C->T | AC | No | NA | NA | NA | NA | intergenic | NA | Yes | Clade Ia, group II to branch 2 excepted KJ642613 | NA |
| 943 | reverse | G->A | GT | No | ACG | T | ATG | M | nonsynonymous | moderately conservative | No | Clade Ia, group II to branch 2 excepted KJ642613 | OPG001 |
| 3344 | reverse | G->T | | No | GCC | A | GCA | A | synonymous | NA | No | Clade Ia, group II to branch 2 excepted KJ642613 | OPG003 |
| 189286 | forward | G->A | GC | No | TGC | C | TAC | Y | nonsynonymous | radical | Yes | Clade Ia, group II to branch 2 excepted KJ642613 | OPG005 |
| 11620 | reverse | G->A | GT | No | ACA | T | ATA | I | nonsynonymous | moderately conservative | No | Clade Ia, group II to branch 2 excepted KJ642613 | OPG023 |
| 33242 | reverse | G->A | GT | No | CTT | L | TTT | F | nonsynonymous | conservative | No | Clade Ia, group II to branch 2 excepted KJ642613 | OPG050 |

| site | direction | snp | dimer | apobec | parent codon | parent aa | child codon | child aa | mutation_category | prediction | homoplasy | Cluster | Remarks | gene-OPG |
|---|---|---|---|---|---|---|---|---|---|---|---|---|---|---|
| 39257 | reverse | G->T | | No | ACC | T | AAC | N | nonsynonymous | moderately conservative | No | Clade Ia, group II to branch 2 excepted KJ642613 | | OPG057 |
| 60179 | reverse | T->C | | No | TCA | S | TCG | S | synonymous | NA | No | Clade Ia, group II to branch 2 excepted KJ642613 | | OPG080 |
| 63257 | reverse | G->A | GT | No | GAC | D | GAT | D | synonymous | NA | No | Clade Ia, group II to branch 2 excepted KJ642613 | | OPG082 |
| 64695 | reverse | G->A | GA | Yes | ATC | I | ATT | I | synonymous | NA | Yes | Clade Ia, group II to branch 2 excepted KJ642613 | | OPG083 |
| 89780 | forward | C->T | GC | No | AGC | S | AGT | S | synonymous | NA | No | Clade Ia, group II to branch 2 excepted KJ642613 | | OPG110 |
| 102930 | forward | C->G | | No | AAC | N | AAG | K | nonsynonymous | moderately conservative | No | Clade Ia, group II to branch 2 excepted KJ642613 | | OPG122 |
| 103393 | reverse | T->C | | No | TCA | S | TCG | S | synonymous | NA | No | Clade Ia, group II to branch 2 excepted KJ642613 | | OPG123 |
| 137852 | reverse | G->T | | No | GCC | A | GCA | A | synonymous | NA | No | Clade Ia, group II to branch 2 excepted KJ642613 | | OPG154 |
| 138900 | reverse | C->T | AC | No | CGT | R | CAT | H | nonsynonymous | conservative | No | Clade Ia, group II to branch 2 excepted KJ642613 | | OPG156 |
| 180253 | forward | G->T | | No | TGA | * | TTA | L | nonsynonymous | NA | No | Clade Ia, group II to branch 2 excepted KJ642613 | | OPG208 |
| 180733 | forward | G->A | GT | No | GTA | V | ATA | I | nonsynonymous | conservative | No | Clade Ia, group II to branch 2 excepted KJ642613 | | OPG209 |
| 32540 | reverse | T->C | | No | GAC | D | GGC | G | nonsynonymous | moderately conservative | No | Clade Ia, group II, branch 2 cluster 1 | | OPG049 |
| 162920 | forward | A->C | | No | CTA | L | CTC | L | synonymous | NA | No | Clade Ia, group II, branch 2 cluster 2 | | OPG188 |

| site | direction | snp | dimer | apobec | parent codon | parent aa | child codon | child aa | mutation_category | prediction | homoplasy | Cluster | Remarks | gene-OPG |
|---|---|---|---|---|---|---|---|---|---|---|---|---|---|---|
| 148732 | reverse | T->G | | No | TAA | * | TAC | Y | nonsynonymous | NA | Yes | Clade Ia, group II, branch 2 , all except cluster 1 | | OPG174 |
| 3734 | reverse | G->A | GA | Yes | ATC | I | ATT | I | synonymous | NA | No | Clade Ia, group II, branch 2 cluster 3 | | OPG003 |
| 21248 | reverse | G->A | GC | No | GGC | G | GGT | G | synonymous | NA | No | Clade Ia, group II, branch 2 cluster 3 | | OPG037 |
| 79586 | forward | T->C | | No | CTA | L | CCA | P | nonsynonymous | moderately conservative | No | Clade Ia, group II, branch 2 cluster 3 | | OPG102 |
| 80959 | reverse | T->C | | No | TCA | S | TCG | S | synonymous | NA | No | Clade Ia, group II, branch 2 cluster 3 | | OPG104 |
| 185841 | forward | A->G | | No | AAC | N | AGC | S | nonsynonymous | conservative | No | Clade Ia, group II, branch 2 cluster 3 | | OPG210 |
| 171 | NA | A->C | | No | NA | NA | NA | NA | intergenic | NA | Yes | Clade Ia, group II, branch 2 cluster 3 to 6 | | NA |
| 26366 | reverse | C->T | TC | Yes | CGA | R | CAA | Q | nonsynonymous | conservative | No | Clade Ia, group II, branch 2 cluster 4 | | OPG042 |
| 54747 | forward | G->T | | No | ATG | M | ATT | I | nonsynonymous | conservative | No | Clade Ia, group II, branch 2 cluster 4 | | OPG072 |
| 64695 | reverse | A->G | | No | ATT | I | ATC | I | synonymous | NA | Yes | Clade Ia, group II, branch 2 cluster 4 | | OPG083 |
| 91902 | forward | G->A | GT | No | CGT | R | CAT | H | nonsynonymous | conservative | No | Clade Ia, group II, branch 2 cluster 4 | | OPG113 |
| 126093 | forward | C->T | GC | No | CAT | H | TAT | Y | nonsynonymous | moderately conservative | No | Clade Ia, group II, branch 2 cluster 4 | | OPG148 |
| 129253 | forward | G->T | | No | GCA | A | TCA | S | nonsynonymous | moderately conservative | No | Clade Ia, group II, branch 2 cluster 4 | | OPG151 |
| 149559 | forward | T->G | | No | GTT | V | GGT | G | nonsynonymous | moderately radical | No | Clade Ia, group II, branch 2 cluster 4 | | OPG175 |
| 151601 | forward | G->A | GT | No | GTG | V | ATG | M | nonsynonymous | conservative | No | Clade Ia, group II, branch 2 cluster 4 | | OPG178 |
| 145464 | NA | T->C | | No | NA | NA | NA | NA | intergenic | NA | No | Clade Ia, group II, branch 2 cluster 5 | | NA |
| 35932 | reverse | C->T | GC | No | GCA | A | ACA | T | nonsynonymous | moderately conservative | No | Clade Ia, group II, branch 2 cluster 5 | | OPG054 |
| 42693 | reverse | G->C | | No | ACA | T | AGA | R | nonsynonymous | moderately conservative | No | Clade Ia, group II, branch 2 cluster 5 | | OPG063 |

| site | direction | snp | dimer | apobec | parent codon | parent aa | child codon | child aa | mutation_category | prediction | homoplasy | Cluster | Remarks | gene-OPG |
|---|---|---|---|---|---|---|---|---|---|---|---|---|---|---|
| 178923 | NA | A->G | | No | NA | NA | NA | NA | intergenic | NA | Yes | Clade Ia, group II, branch 2 cluster 6 | | NA |
| 159653 | forward | A->T | | No | TTA | L | TTT | F | nonsynonymous | conservative | Yes | Clade Ia, group II, branch 2 cluster 6 | | OPG185 |
| 826 | NA | C->T | TC | Yes | NA | NA | NA | NA | intergenic | NA | No | Clade Ia, group III | | NA |
| 157592 | NA | C->A | | No | NA | NA | NA | NA | intergenic | NA | No | Clade Ia, group III | | NA |
| 168929 | NA | G->T | | No | NA | NA | NA | NA | intergenic | NA | Yes | Clade Ia, group III | | NA |
| 178923 | NA | A->G | | No | NA | NA | NA | NA | intergenic | NA | Yes | Clade Ia, group III | | NA |
| 118195 | reverse | C->T | AC | No | TAG | * | TAA | * | synonymous | NA | No | Clade Ia, group III | | OPG136 |
| 150201 | forward | A->T | | No | AAA | K | TAA | * | nonsense | NA | Yes | Clade Ia, group III | | OPG176 |
| 149978 | forward | G->A | GT | No | ATG | M | ATA | I | nonsynonymous | conservative | Yes | Clade Ia, group III | | OPG176 |
| 154113 | forward | G->A | GT | No | GTT | V | ATT | I | nonsynonymous | conservative | No | Clade Ia, group III | | OPG180 |
| 176367 | forward | C->T | AC | No | ACA | T | ATA | I | nonsynonymous | moderately conservative | No | Clade Ia, group III | | OPG205 |
| 177394 | forward | C->T | TC | Yes | GTC | V | GTT | V | synonymous | NA | No | Clade Ia, group III | | OPG205 |
| 186579 | forward | G->T | | No | AGA | R | ATA | I | nonsynonymous | moderately conservative | No | Clade Ia, group III | | OPG210 |
| 168929 | NA | G->T | | No | NA | NA | NA | NA | intergenic | NA | Yes | Clade Ia, novel cluster closed to group III | | NA |
| 23448 | reverse | A->G | | No | TAC | Y | CAC | H | nonsynonymous | moderately conservative | No | Clade Ia, novel cluster closed to group III | | OPG039 |
| 48566 | forward | G->A | GC | No | GCA | A | ACA | T | nonsynonymous | moderately conservative | Yes | Clade Ia, novel cluster closed to group III | | OPG068 |
| 59282 | reverse | G->A | GT | No | CAC | H | CAT | H | synonymous | NA | No | Clade Ia, novel cluster closed to group III | | OPG079 |
| 65664 | forward | C->T | AC | No | ACG | T | ATG | M | nonsynonymous | moderately conservative | No | Clade Ia, novel cluster closed to group III | | OPG084 |
| 86071 | forward | G->A | GC | No | ACG | T | ACA | T | synonymous | NA | No | Clade Ia, novel cluster closed to group III | | OPG107 |
| 113943 | reverse | C->T | GC | No | GCC | A | ACC | T | nonsynonymous | moderately conservative | No | Clade Ia, novel cluster closed to group III | | OPG133 |
| 140728 | reverse | G->A | GA | Yes | CAT | H | TAT | Y | nonsynonymous | moderately conservative | No | Clade Ia, novel cluster closed to group III | | OPG160 |
| 147889 | forward | A->C | | No | AAT | N | ACT | T | nonsynonymous | moderately conservative | No | Clade Ia, novel cluster closed to group III | | OPG172 |

| site | direction | snp | dimer | apobec | parent codon | parent aa | child codon | child aa | mutation_category | prediction | homoplasy | Cluster Remarks | gene-OPG |
|---|---|---|---|---|---|---|---|---|---|---|---|---|---|
| 177066 | forward | C->T | GC | No | GCT | A | GTT | V | nonsynonymous | moderately conservative | No | Clade Ia, novel cluster closed to group III | OPG205 |
| 185470 | forward | G->A | GA | Yes | GTG | V | GTA | V | synonymous | NA | No | Clade Ia, novel cluster closed to group III | OPG210 |
| 19049 | NA | T->A | | No | NA | NA | NA | NA | intergenic | NA | No | Clade Ia, group IV | NA |
| 89540 | NA | C->T | GC | No | NA | NA | NA | NA | intergenic | NA | No | Clade Ia, group IV | NA |
| 148301 | NA | G->A | GG | No | NA | NA | NA | NA | intergenic | NA | No | Clade Ia, group IV | NA |
| 42209 | forward | G->A | GC | No | ATG | M | ATA | I | nonsynonymous | conservative | No | Clade Ia, group IV | OPG062 |
| 56842 | reverse | C->A | | No | GTT | V | TTT | F | nonsynonymous | conservative | No | Clade Ia, group IV | OPG074 |
| 58670 | reverse | G->T | | No | ACC | T | ACA | T | synonymous | NA | No | Clade Ia, group IV | OPG078 |
| 59951 | reverse | C->T | GC | No | GTG | V | GTA | V | synonymous | NA | No | Clade Ia, group IV | OPG080 |
| 84253 | forward | C->A | | No | CCC | P | CCA | P | synonymous | NA | No | Clade Ia, group IV | OPG105 |
| 111818 | reverse | C->A | | No | CCG | P | CCT | P | synonymous | NA | No | Clade Ia, group IV | OPG130 |
| 124688 | forward | G->A | GC | No | CGC | R | CAC | H | nonsynonymous | conservative | Yes | Clade Ia, group IV | OPG145 |
| 136139 | reverse | A->G | | No | TAG | * | CAG | Q | nonsynonymous | NA | No | Clade Ia, group IV | OPG153 |
| 147805 | forward | G->A | GC | No | TGC | C | TAC | Y | nonsynonymous | radical | No | Clade Ia, group IV | OPG172 |
| 164819 | forward | A->G | | No | ATA | I | ATG | M | nonsynonymous | conservative | No | Clade Ia, group IV | OPG189 |
| 186992 | forward | G->A | GA | Yes | GAA | E | AAA | K | nonsynonymous | moderately conservative | Yes | Clade Ia, group IV | OPG210 |
| 25699 | NA | G->T | | No | NA | NA | NA | NA | intergenic | NA | No | Clade Ia, group V (branch to JX878417) | NA |
| 33749 | NA | G->A | GC | No | NA | NA | NA | NA | intergenic | NA | Yes | Clade Ia, group V (branch to JX878417) | NA |
| 49955 | NA | C->T | GC | No | NA | NA | NA | NA | intergenic | NA | No | Clade Ia, group V (branch to JX878417) | NA |
| 158859 | NA | C->T | AC | No | NA | NA | NA | NA | intergenic | NA | No | Clade Ia, group V (branch to JX878417) | NA |
| 169483 | NA | C->T | AC | No | NA | NA | NA | NA | intergenic | NA | Yes | Clade Ia, group V (branch to JX878417) | NA |
| 178923 | NA | A->G | | No | NA | NA | NA | NA | intergenic | NA | Yes | Clade Ia, group V (branch to JX878417) | NA |
| 178986 | NA | C->T | TC | Yes | NA | NA | NA | NA | intergenic | NA | No | Clade Ia, group V (branch to JX878417) | NA |

| site | direction | snp | dimer | apobec | parent codon | parent aa | child codon | child aa | mutation_category | prediction | homoplasy | Cluster | Remarks | gene-OPG |
|---|---|---|---|---|---|---|---|---|---|---|---|---|---|---|
| 180434 | NA | T->A | | No | NA | NA | NA | NA | intergenic | NA | No | Clade Ia, group V (branch to JX878417) | | NA |
| 189289 | forward | G->A | GC | No | TGC | C | TAC | Y | nonsynonymous | radical | No | Clade Ia, group V (branch to JX878417) | | OPG005 |
| 16202 | reverse | G->A | GT | No | CAC | H | TAC | Y | nonsynonymous | moderately conservative | No | Clade Ia, group V (branch to JX878417) | | OPG027 |
| 19836 | reverse | A->C | | No | TAA | * | GAA | E | nonsynonymous | NA | No | Clade Ia, group V (branch to JX878417) | | OPG034 |
| 19432 | reverse | G->A | GC | No | GGC | G | GGT | G | synonymous | NA | No | Clade Ia, group V (branch to JX878417) | | OPG034 |
| 31836 | reverse | T->C | | No | ATA | I | GTA | V | nonsynonymous | conservative | No | Clade Ia, group V (branch to JX878417) | | OPG048 |
| 41758 | reverse | C->T | GC | No | CCG | P | CCA | P | synonymous | NA | No | Clade Ia, group V (branch to JX878417) | | OPG061 |
| 49318 | forward | G->T | | No | ATG | M | ATT | I | nonsynonymous | conservative | No | Clade Ia, group V (branch to JX878417) | | OPG068 |
| 51487 | reverse | A->G | | No | TTC | F | TCC | S | nonsynonymous | radical | No | Clade Ia, group V (branch to JX878417) | | OPG071 |
| 62009 | reverse | G->A | GC | No | TGC | C | TGT | C | synonymous | NA | No | Clade Ia, group V (branch to JX878417) | | OPG080 |
| 62183 | reverse | A->G | | No | AGT | S | AGC | S | synonymous | NA | No | Clade Ia, group V (branch to JX878417) | | OPG081 |
| 72640 | reverse | T->C | | No | ATC | I | GTC | V | nonsynonymous | conservative | No | Clade Ia, group V (branch to JX878417) | | OPG092 |
| 73232 | forward | T->G | | No | TCT | S | GCT | A | nonsynonymous | moderately conservative | No | Clade Ia, group V (branch to JX878417) | | OPG093 |
| 75313 | forward | C->T | GC | No | GCG | A | GTG | V | nonsynonymous | moderately conservative | No | Clade Ia, group V (branch to JX878417) | | OPG095 |
| 89371 | reverse | G->T | | No | CGT | R | AGT | S | nonsynonymous | moderately radical | No | Clade Ia, group V (branch to JX878417) | | OPG109 |
| 93791 | forward | C->T | GC | No | CGT | R | TGT | C | nonsynonymous | radical | No | Clade Ia, group V (branch to JX878417) | | OPG113 |
| 94019 | forward | G->A | GC | No | GCC | A | ACC | T | nonsynonymous | moderately conservative | Yes | Clade Ia, group V (branch to JX878417) | | OPG113 |
| 112442 | reverse | C->A | | No | ACG | T | ACT | T | synonymous | NA | No | Clade Ia, group V (branch to JX878417) | | OPG132 |

| site | direction | snp | dimer | apobec | parent codon | parent aa | child codon | child aa | mutation_category | prediction | homoplasy | Cluster | Remarks | gene-OPG |
|---|---|---|---|---|---|---|---|---|---|---|---|---|---|---|
| 119429 | reverse | G->A | GT | No | ACT | T | ATT | I | nonsynonymous | moderately conservative | No | Clade Ia, group V (branch to JX878417) | | OPG136 |
| 124688 | forward | G->A | GC | No | CGC | R | CAC | H | nonsynonymous | conservative | Yes | Clade Ia, group V (branch to JX878417) | | OPG145 |
| 144147 | forward | A->G | | No | GTA | V | GTG | V | synonymous | NA | No | Clade Ia, group V (branch to JX878417) | | OPG165 |
| 148082 | forward | C->T | CC | No | CCG | P | CTG | L | nonsynonymous | moderately conservative | No | Clade Ia, group V (branch to JX878417) | | OPG173 |
| 150182 | forward | C->A | | No | ACC | T | ACA | T | synonymous | NA | No | Clade Ia, group V (branch to JX878417) | | OPG176 |
| 153019 | forward | T->C | | No | GTA | V | GCA | A | nonsynonymous | moderately conservative | No | Clade Ia, group V (branch to JX878417) | | OPG180 |
| 168022 | forward | C->T | GC | No | TGC | C | TGT | C | synonymous | NA | No | Clade Ia, group V (branch to JX878417) | | OPG193 |
| 175035 | forward | C->T | AC | No | CGT | R | TGT | C | nonsynonymous | radical | No | Clade Ia, group V (branch to JX878417) | | OPG204 |
| 180920 | forward | G->A | GC | No | AGC | S | AAC | N | nonsynonymous | conservative | Yes | Clade Ia, group V (branch to JX878417) | | OPG209 |
| 7024 | NA | C->T | AC | No | NA | NA | NA | NA | intergenic | NA | No | Clade Ib | | NA |
| 8042 | NA | C->T | GC | No | NA | NA | NA | NA | intergenic | NA | No | Clade Ib | | NA |
| 8743 | NA | C->T | GC | No | NA | NA | NA | NA | intergenic | NA | No | Clade Ib | | NA |
| 16560 | NA | C->T | CC | No | NA | NA | NA | NA | intergenic | NA | No | Clade Ib | | NA |
| 18958 | NA | C->T | GC | No | NA | NA | NA | NA | intergenic | NA | No | Clade Ib | | NA |
| 19038 | NA | A->C | | No | NA | NA | NA | NA | intergenic | NA | No | Clade Ib | | NA |
| 19128 | NA | T->A | | No | NA | NA | NA | NA | intergenic | NA | No | Clade Ib | | NA |
| 20453 | NA | T->C | | No | NA | NA | NA | NA | intergenic | NA | No | Clade Ib | | NA |
| 20455 | NA | C->A | | No | NA | NA | NA | NA | intergenic | NA | No | Clade Ib | | NA |
| 20456 | NA | A->T | | No | NA | NA | NA | NA | intergenic | NA | No | Clade Ib | | NA |
| 25635 | NA | C->A | | No | NA | NA | NA | NA | intergenic | NA | No | Clade Ib | | NA |
| 47658 | NA | G->A | GT | No | NA | NA | NA | NA | intergenic | NA | Yes | Clade Ib | | NA |
| 81085 | NA | C->T | TC | Yes | NA | NA | NA | NA | intergenic | NA | No | Clade Ib | | NA |
| 133915 | NA | G->A | GT | No | NA | NA | NA | NA | intergenic | NA | No | Clade Ib | | NA |
| 133936 | NA | A->G | | No | NA | NA | NA | NA | intergenic | NA | Yes | Clade Ib | | NA |

| site | direction | snp | dimer | apobec | parent codon | parent aa | child codon | child aa | mutation_category | prediction | homoplasy | Cluster | Remarks | gene-OPG |
|---|---|---|---|---|---|---|---|---|---|---|---|---|---|---|
| 135422 | NA | G->A | GT | No | NA | NA | NA | NA | intergenic | NA | No | Clade Ib | | NA |
| 156500 | NA | C->T | GC | No | NA | NA | NA | NA | intergenic | NA | Yes | Clade Ib | | NA |
| 156635 | NA | C->T | AC | No | NA | NA | NA | NA | intergenic | NA | Yes | Clade Ib | | NA |
| 158612 | NA | C->T | AC | No | NA | NA | NA | NA | intergenic | NA | No | Clade Ib | | NA |
| 160116 | NA | G->T | | No | NA | NA | NA | NA | intergenic | NA | No | Clade Ib | | NA |
| 165027 | NA | C->T | GC | No | NA | NA | NA | NA | intergenic | NA | Yes | Clade Ib | | NA |
| 170928 | NA | C->A | | No | NA | NA | NA | NA | intergenic | NA | No | Clade Ib | | NA |
| 178130 | NA | G->T | | No | NA | NA | NA | NA | intergenic | NA | No | Clade Ib | | NA |
| 3181 | reverse | G->A | GT | No | CCA | P | TCA | S | nonsynonymous | moderately conservative | No | Clade Ib | | OPG003 |
| 3003 | reverse | G->A | GG | No | CCA | P | CTA | L | nonsynonymous | moderately conservative | No | Clade Ib | | OPG003 |
| 5366 | reverse | C->T | GC | No | GCA | A | ACA | T | nonsynonymous | moderately conservative | No | Clade Ib | | OPG015 |
| 5968 | reverse | A->G | | No | ATA | I | ACA | T | nonsynonymous | moderately conservative | No | Clade Ib | | OPG015 |
| 7889 | forward | G->T | | No | AGG | R | ATG | M | nonsynonymous | moderately conservative | No | Clade Ib | | OPG019 |
| 12625 | reverse | G->A | GC | No | GCT | A | GTT | V | nonsynonymous | moderately conservative | No | Clade Ib | | OPG023 |
| 14498 | reverse | G->T | | No | CCA | P | ACA | T | nonsynonymous | conservative | No | Clade Ib | | OPG025 |
| 20269 | reverse | G->A | GA | Yes | ATC | I | ATT | I | synonymous | NA | No | Clade Ib | | OPG035 |
| 21460 | reverse | G->T | | No | CTA | L | ATA | I | nonsynonymous | conservative | No | Clade Ib | | OPG037 |
| 23021 | reverse | C->T | AC | No | TGG | W | TGA | * | nonsense | NA | No | Clade Ib | | OPG038 |
| 27032 | reverse | C->T | GC | No | TGC | C | TAC | Y | nonsynonymous | radical | No | Clade Ib | | OPG042 |
| 28658 | reverse | G->A | GT | No | CTC | L | TTC | F | nonsynonymous | conservative | No | Clade Ib | | OPG045 |
| 29788 | reverse | T->C | | No | AAT | N | AGT | S | nonsynonymous | conservative | No | Clade Ib | | OPG047 |
| 33551 | reverse | C->T | TC | Yes | CGA | R | CAA | Q | nonsynonymous | conservative | No | Clade Ib | | OPG051 |
| 43441 | reverse | T->C | | No | ATT | I | GTT | V | nonsynonymous | conservative | No | Clade Ib | | OPG063 |
| 43996 | reverse | A->G | | No | TGT | C | TGC | C | synonymous | NA | No | Clade Ib | | OPG064 |
| 44254 | reverse | C->T | AC | No | ACG | T | ACA | T | synonymous | NA | No | Clade Ib | | OPG064 |

| site | direction | snp | dimer | apobec | parent codon | parent aa | child codon | child aa | mutation_category | prediction | homoplasy | Cluster | Remarks | gene-OPG |
|---|---|---|---|---|---|---|---|---|---|---|---|---|---|---|
| 48612 | forward | A->T | | No | AAT | N | ATT | I | nonsynonymous | moderately radical | No | Clade Ib | | OPG068 |
| 48786 | forward | G->T | | No | AGA | R | ATA | I | nonsynonymous | moderately conservative | No | Clade Ib | | OPG068 |
| 49030 | forward | C->A | | No | TCC | S | TCA | S | synonymous | NA | Yes | Clade Ib | | OPG068 |
| 52895 | reverse | T->C | | No | ACA | T | GCA | A | nonsynonymous | moderately conservative | No | Clade Ib | | OPG071 |
| 54708 | forward | A->G | | No | CCA | P | CCG | P | synonymous | NA | No | Clade Ib | | OPG072 |
| 64899 | forward | T->A | | No | CTA | L | CAA | Q | nonsynonymous | moderately radical | No | Clade Ib | | OPG084 |
| 69405 | forward | C->T | TC | Yes | CAA | Q | TAA | * | nonsense | NA | No | Clade Ib | | OPG087 |
| 78083 | forward | T->C | | No | CTA | L | CCA | P | nonsynonymous | moderately conservative | Yes | Clade Ib | | OPG100 |
| 78255 | forward | G->A | GG | No | TCG | S | TCA | S | synonymous | NA | No | Clade Ib | | OPG100 |
| 79916 | forward | G->A | GA | Yes | CGA | R | CAA | Q | nonsynonymous | conservative | No | Clade Ib | | OPG102 |
| 80681 | reverse | G->A | GT | No | ACG | T | ATG | M | nonsynonymous | moderately conservative | No | Clade Ib | | OPG104 |
| 88616 | reverse | G->A | GT | No | CAC | H | CAT | H | synonymous | NA | No | Clade Ib | | OPG109 |
| 91583 | forward | G->A | GT | No | CGT | R | CAT | H | nonsynonymous | conservative | No | Clade Ib | | OPG112 |
| 108459 | reverse | G->A | GT | No | TAC | Y | TAT | Y | synonymous | NA | No | Clade Ib | | OPG127 |
| 110338 | reverse | C->A | | No | AAG | K | AAT | N | nonsynonymous | moderately conservative | No | Clade Ib | | OPG129 |
| 111091 | reverse | A->G | | No | TAG | * | CAG | Q | nonsynonymous | NA | No | Clade Ib | | OPG130 |
| 111249 | reverse | C->T | CC | No | CGG | R | CAG | Q | nonsynonymous | conservative | No | Clade Ib | | OPG130 |
| 119651 | forward | C->T | GC | No | AGC | S | AGT | S | synonymous | NA | No | Clade Ib | | OPG137 |
| 120932 | reverse | G->A | GT | No | ACA | T | ATA | I | nonsynonymous | moderately conservative | No | Clade Ib | | OPG138 |
| 120962 | reverse | C->T | CC | No | TGG | W | TAG | * | nonsense | NA | No | Clade Ib | | OPG138 |
| 123914 | reverse | G->T | | No | TCC | S | TCA | S | synonymous | NA | No | Clade Ib | | OPG144 |
| 123987 | forward | G->A | GC | No | TAG | * | TAA | * | synonymous | NA | No | Clade Ib | | OPG145 |
| 126456 | forward | C->A | | No | CAA | Q | AAA | K | nonsynonymous | moderately conservative | No | Clade Ib | | OPG148 |
| 132286 | forward | A->G | | No | AAC | N | GAC | D | nonsynonymous | conservative | No | Clade Ib | | OPG151 |

| site | direction | snp | dimer | apobec | parent codon | parent aa | child codon | child aa | mutation_category | prediction | homoplasy | Cluster | Remarks | gene-OPG |
|---|---|---|---|---|---|---|---|---|---|---|---|---|---|---|
| 132024 | forward | G->A | GA | Yes | ACG | T | ACA | T | synonymous | NA | No | Clade Ib | | OPG151 |
| 136425 | reverse | G->A | GT | No | GAC | D | GAT | D | synonymous | NA | No | Clade Ib | | OPG153 |
| 145002 | reverse | A->T | | No | TAT | Y | TAA | * | nonsense | NA | No | Clade Ib | | OPG167 |
| 148361 | reverse | C->T | AC | No | TGT | C | TAT | Y | nonsynonymous | radical | No | Clade Ib | | OPG174 |
| 149955 | forward | A->G | | No | ACT | T | GCT | A | nonsynonymous | moderately conservative | No | Clade Ib | | OPG176 |
| 150310 | forward | G->A | GC | No | CGC | R | CAC | H | nonsynonymous | conservative | No | Clade Ib | | OPG176 |
| 150200 | forward | T->A | | No | TTT | F | TTA | L | nonsynonymous | conservative | Yes | Clade Ib | | OPG176 |
| 154518 | forward | T->C | | No | TGT | C | TGC | C | synonymous | NA | No | Clade Ib | | OPG181 |
| 159332 | forward | C->T | AC | No | AAC | N | AAT | N | synonymous | NA | No | Clade Ib | | OPG185 |
| 162942 | forward | A->C | | No | ATA | I | CTA | L | nonsynonymous | conservative | No | Clade Ib | | OPG188 |
| 182469 | forward | C->T | GC | No | GCG | A | GTG | V | nonsynonymous | moderately conservative | No | Clade Ib | | OPG210 |
| 184213 | forward | G->T | | No | GAG | E | GAT | D | nonsynonymous | conservative | No | Clade Ib | | OPG210 |
