## Supplementary figures and images for "Clade I Mpox virus genomic diversity in the Democratic Republic of the Congo, 2018 - 2024: Predominance of Zoonotic Transmission"

### Supplementary figure 1

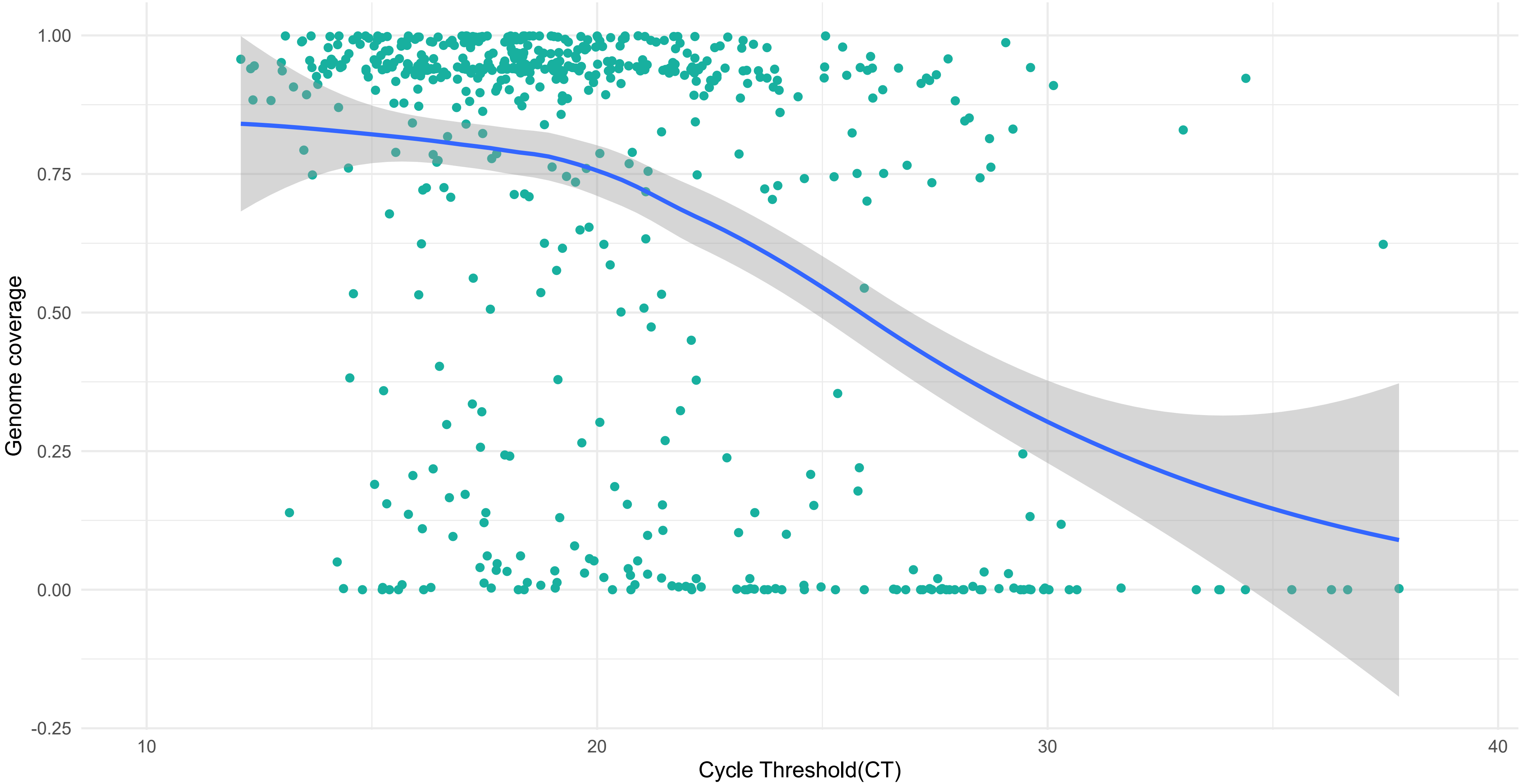


**Figure S1** :

**Scatterplot of genome called coverage vs PCR Cycle threshold**

### Supplementary figure 2

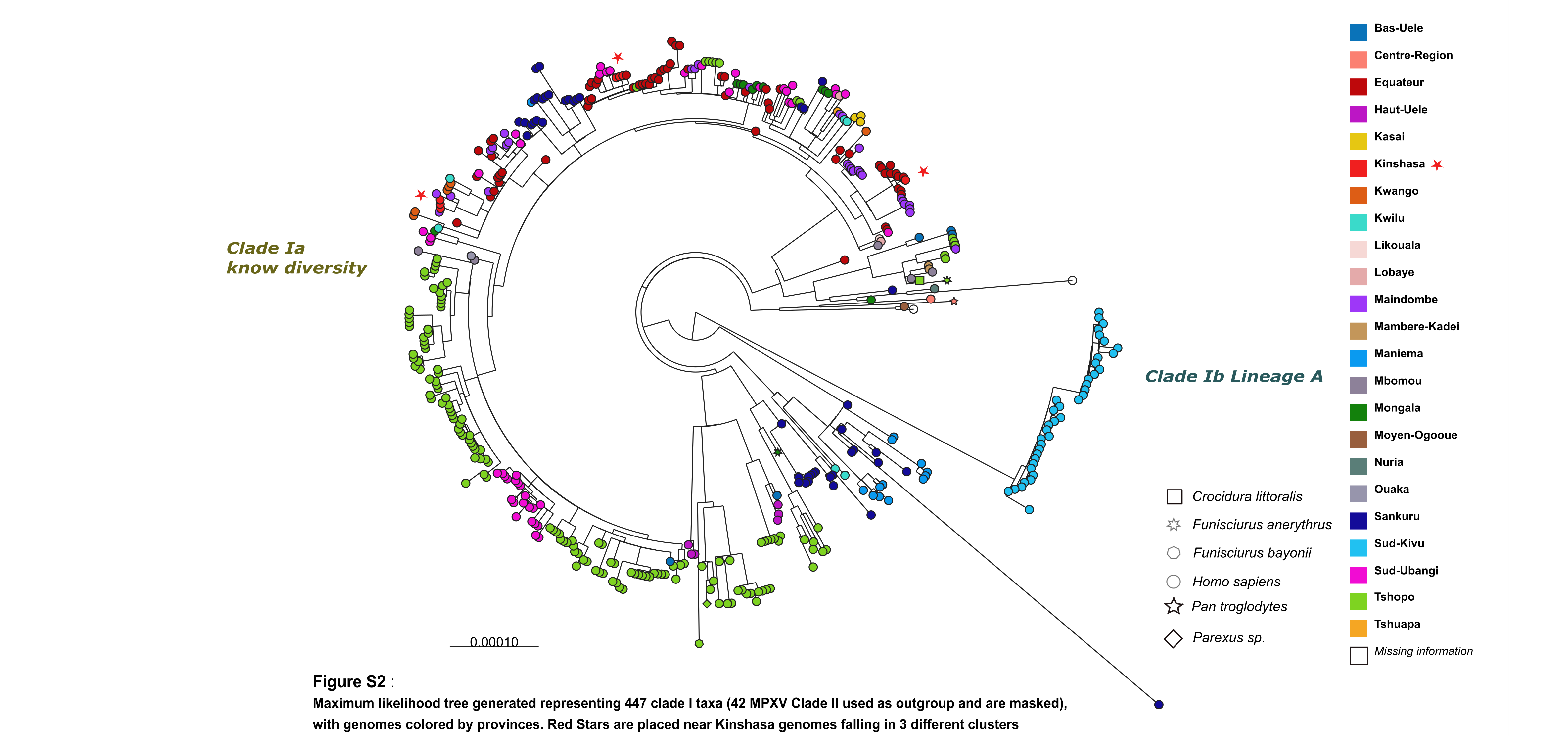
