## Supplementary figure 3 for "Clade I Mpox virus genomic diversity in the Democratic Republic of the Congo, 2018 - 2024: Predominance of Zoonotic Transmission"

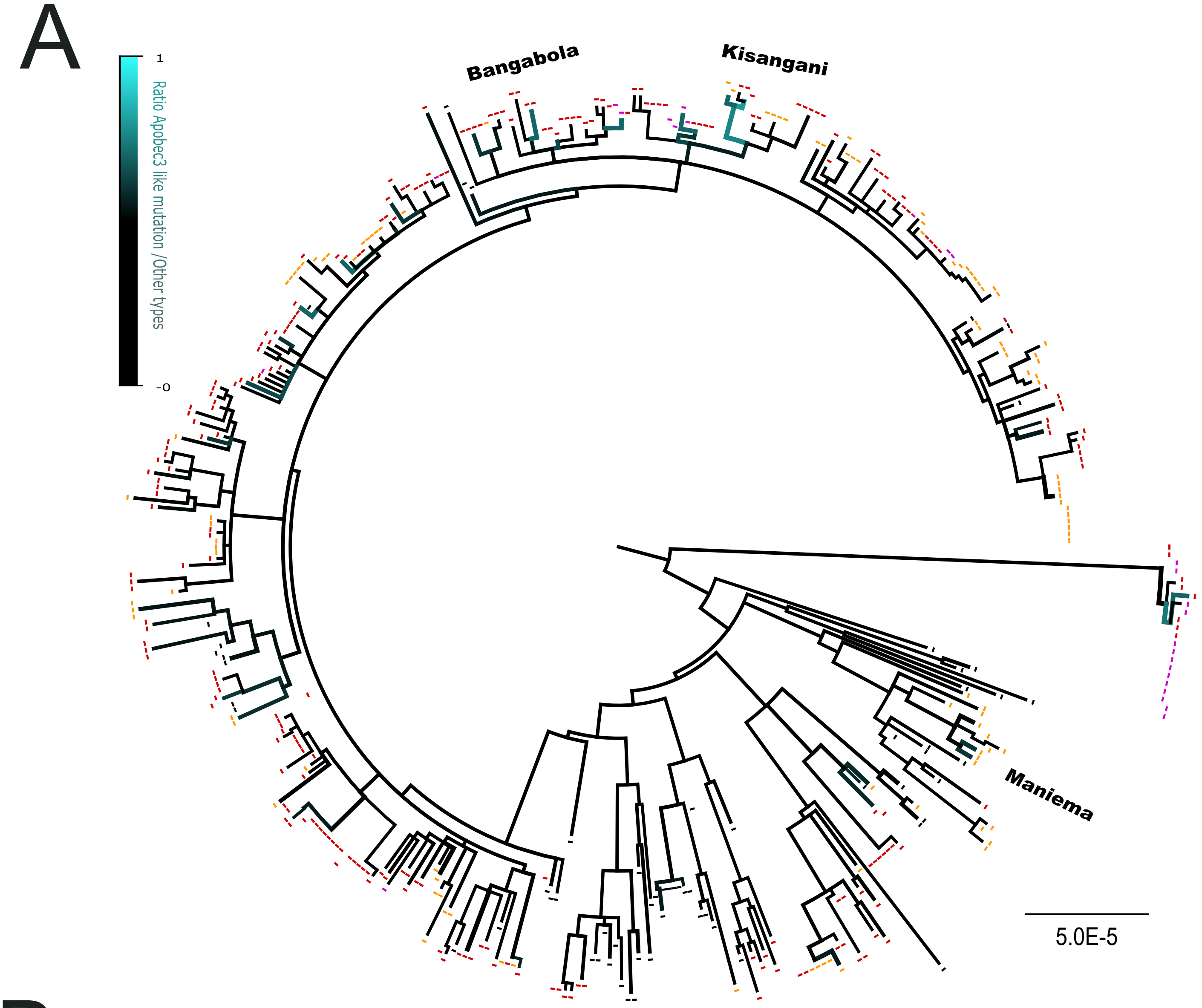


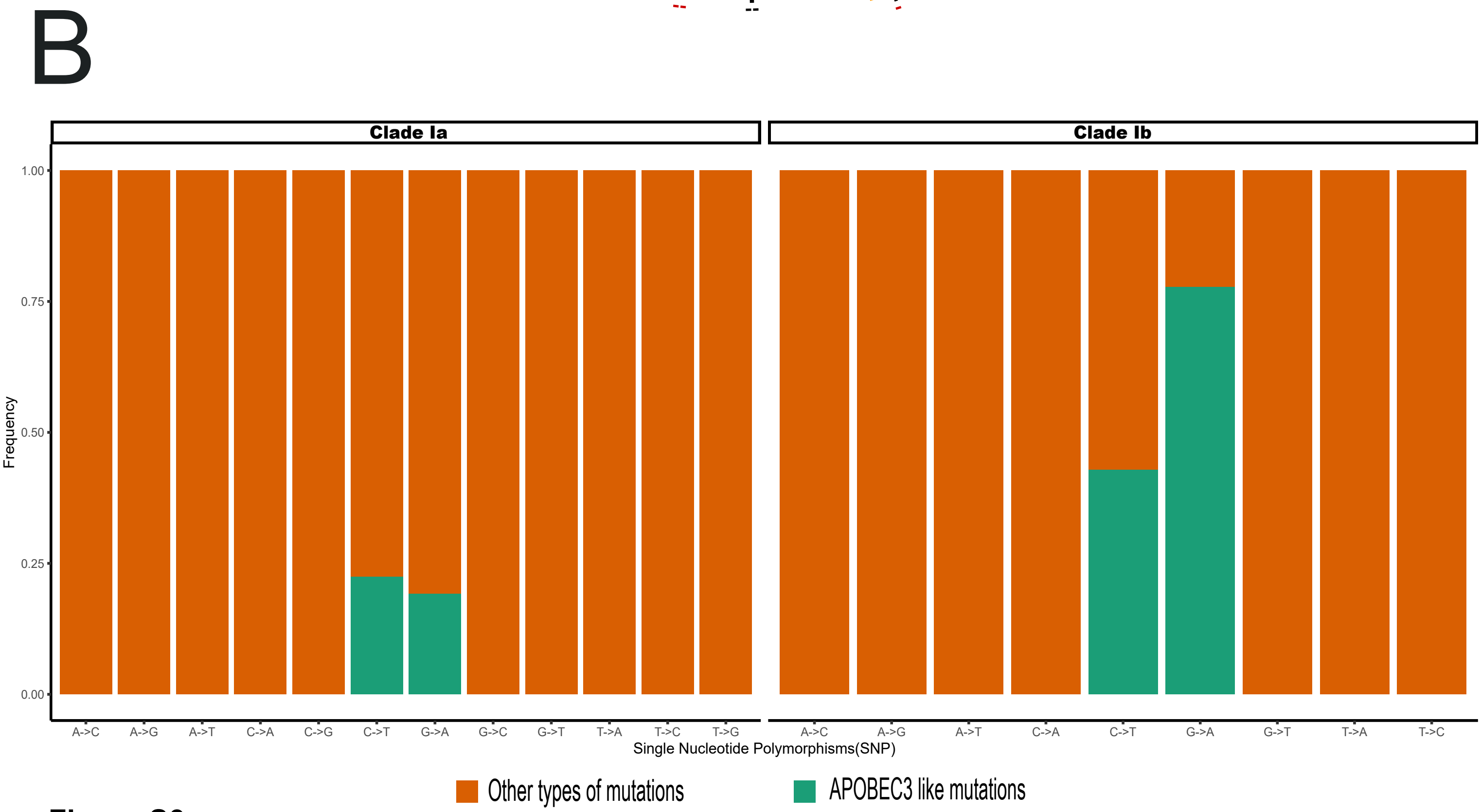


**Figure S3** :

**(A)Apobec3 analysis box at tips are colored by the year (black = prior 2022, orange = 2022, red = 2023, purple = 2024) and branches width and color gradient to cyan are consistent with proportion apobec3 mutations/others-**

**(B) Proportion of Single Nucleotide Polymorphism and apobec like mutations per subclade within Clade I**
